# Molecular Insights into β-Lactams Resistance in *Klebsiella pneumoniae* Clinical Isolates with a Focus on Multidrug Resistance and Virulence

**DOI:** 10.1101/2023.12.30.23300671

**Authors:** Lavouisier F.B. Nogueira, Marília S. Maia, Marco A.F. Clementino, Ila F.N. Lima, Jorge L.N. Rodrigues, Luciana V.C. Fragoso, Glairta S. Costa, Jose Q.S. Filho, Alexandre Havt, Deiziane V.S. Costa, Lyvia M.V.C. Magalhães, Dilza Silva, Nicholas E. Sherman, José K. Sousa, Aldo A.M. Lima

## Abstract

*Klebsiella pneumoniae* is associated with high resistance to antimicrobials and is common in isolates from colonization and healthcare-associated infections (HAIs). This study aims to develop assays to detect resistance genes belonging to the *bla* family and investigate metabolic pathways in resistant isolates of *K. pneumoniae*. The genes of the subfamilies included were: *bla*SHV*, bla* TEM*, bla*NDM*, bla*KPC*, bla* GES*, bla* CTX-M and relevant variants of the *bla*OXA subfamily. Mass spectrometry data were acquired on the Orbitrap IDX spectrometer (Thermo) connected to the Vanquish UPLC system. Isolates of *K. pneumoniae* (N = 122) were obtained from clinical samples from 04/23/2019 to 05/29/2021. A high prevalence of resistance to penicillins, cephalosporins and carbapenems was found among the isolates. The identified genotypic profile showed a high prevalence of genes belonging to Ambler’s classes of beta-lactamases A, B and D. In the metabolomic study, the N-fructosyl isoleucine metabolite was identified increased in multidrug-resistant (MDR) strains of *K. pneumoniae* compared to strains susceptible to antimicrobials. In conclusion, the assays developed were efficient in detecting the main genes of the *bla* family of resistance in *K. pneumoniae*. The use of the pentose phosphate metabolic pathway suggests a beneficial regulation of bacterial growth, and colonization in MDR *K. pneumoniae* strains, with may indicate the use of this pathway as a virulence mechanism in resistant strains.

## INTRODUCTION

Bacterial resistance to antimicrobial agents is a global public health problem that leads to an increase in the cost of treatment, length of stay, morbidity and mortality of hospitalized patients, especially in intensive care units (ICU).^1,2^ In this sense, understanding the emergency mechanisms of regulation and dissemination of resistance to antimicrobial agents in the pathogenesis of bacterial colonization and/or infection can be of critical importance in preventing and controlling this problem.

*Klebsiella pneumoniae* is a Gram-negative γ-proteobacteria bacteria, belonging to the *Enterobacteriacae* family, and is generally viewed as an opportunistic microorganism, carrying several virulence factors and capable of accumulating resistance genes to various classes of antimicrobials. It is commonly related to cases of colonization and/or healthcare-associated infections (HAIs) and has been identified as an etiological agent in pneumonia, urinary tract infections (UTI), soft tissue and surgical wound infections, bacteremia, and sepsis.^3^

It is estimated that *K. pneumoniae* is responsible for approximately 10% of all cases of HAIs, and of these, 32.8% are caused by strains resistant to multiple antimicrobial drugs. However, studies indicate that the rate of isolated strains presenting resistance to antimicrobials has increased over the years.^4,5^

*K. pneumoniae* has been associated with the ability to overcome colonization resistance imposed by the gastrointestinal microbiota^6^ and has been reported as an emerging Multidrug-Resistant microorganism of emergency priority by the World Health Organization (WHO) for the development of new therapies.^7^ Epidemiological data have demonstrated that *K. pneumoniae* can translocate from the gastrointestinal tract to other sterile sites of the same host or other patients through the fecal-oral route. What makes clear the clinical relevance of this microorganism. ^8^

One of the most likely causes of the increasingly frequent emergence of bacterial strains resistant to one or more antibiotics is the excessive and sometimes incorrect use of antimicrobials. The relatively long time required to identify the pathogen by traditional methods, as well as for the results of the antimicrobial susceptibility test (AST), forces the clinician to use broad-spectrum drugs empirically, increasing selective pressure, which ends up benefiting pathogens genetically capable of adapting to the adverse environment.^1,9^

Possible answers to reduce the time needed to obtain a resistance profile, and to investigate new approaches in treatment, is the development of molecular methodologies, that can make the identification of resistance faster, and also be more sensitive than the traditional methodologies.^10^ And the investigation of metabolic differences between sensitive and resistant strains, that can lead to metabolites or pathways that can be used as targets for the development of new treatments and strategies for dealing with multidrug-resistant (MDR) microorganisms. ^11–12^

Beta-lactams are the class of antimicrobials most affected by resistance in general, which is conferred in gram-negatives, mainly by the *bla* gene family, this group of genes encodes enzymes called beta-lactamases, which can neutralize the action of beta-lactams through hydrolysis of the beta-lactam ring.^13–15^

In this study it was investigated the phenotype of resistance, the genotypic profile of the *bla* family of β-lactamases, and the metabolic differences between sensitive and MDR *K. pneumoniae* strains, isolated from patients admitted to the ICU in a university hospital in the city of Fortaleza-CE, Brazil.

We aim to develop a molecular assay, capable of identifying all variants of the most relevant genes of the *bla* family, these being: *bla*SHV*, bla*TEM*, bla*NDM*, bla*KPC*, bla*GES and *b l* C*a* TX-M. In addition, also was included the most epidemiologically relevant variants of the *bla-*OXA subfamily. Thus, creating a set of primers capable of detecting the presence of hundreds of resistance genes with a reduced number of reactions. Were also investigated the main differences between the metabolites up and down regulated in sensitive and MDR strains of *K. pneumoniae*, both from the intracellular medium and the supernatant among themselves and in comparison, with the control medium.

## RESULTS

### Selection of bacterial samples

A total of 249 samples of Gram-negative bacteria resistant to beta lactam antimicrobials were collected from 04/23/2019 to 05/29/2021, those were identified, and the most prevalent microorganism found was *Klebsiella pneumoniae* 48.99% (122/249), been 21,8% from infections cases and 78,2% from colonization cases.

Identification of the phenotypic profile of beta-lactams resistance of K. pneumoniae strains. The bacteria isolates included in the study were tested against a wide range of beta-lactams drugs, and the presence of a high percentage of isolates resistant to these drugs was verified, which included <u>Penicillins</u>: ampicillin/sulbactam (92.00%) and piperacillin/tazobactam (88.46%), <u>Cephalosporins</u>: cefepime (83.02%), cefoxitin (73.68%), ceftazidime (86.54%), ceftazidime/avibactam (11 .76%), ceftriaxone (83.02%), cefuroxime (90.38%) and cefuroxime axetil (81.82%) and <u>Carbapenems</u>: ertapenem (44.00%), imipenem (71.70%) and meropenem (69.81%), as shown in Figure 1.

**Figure 1.**
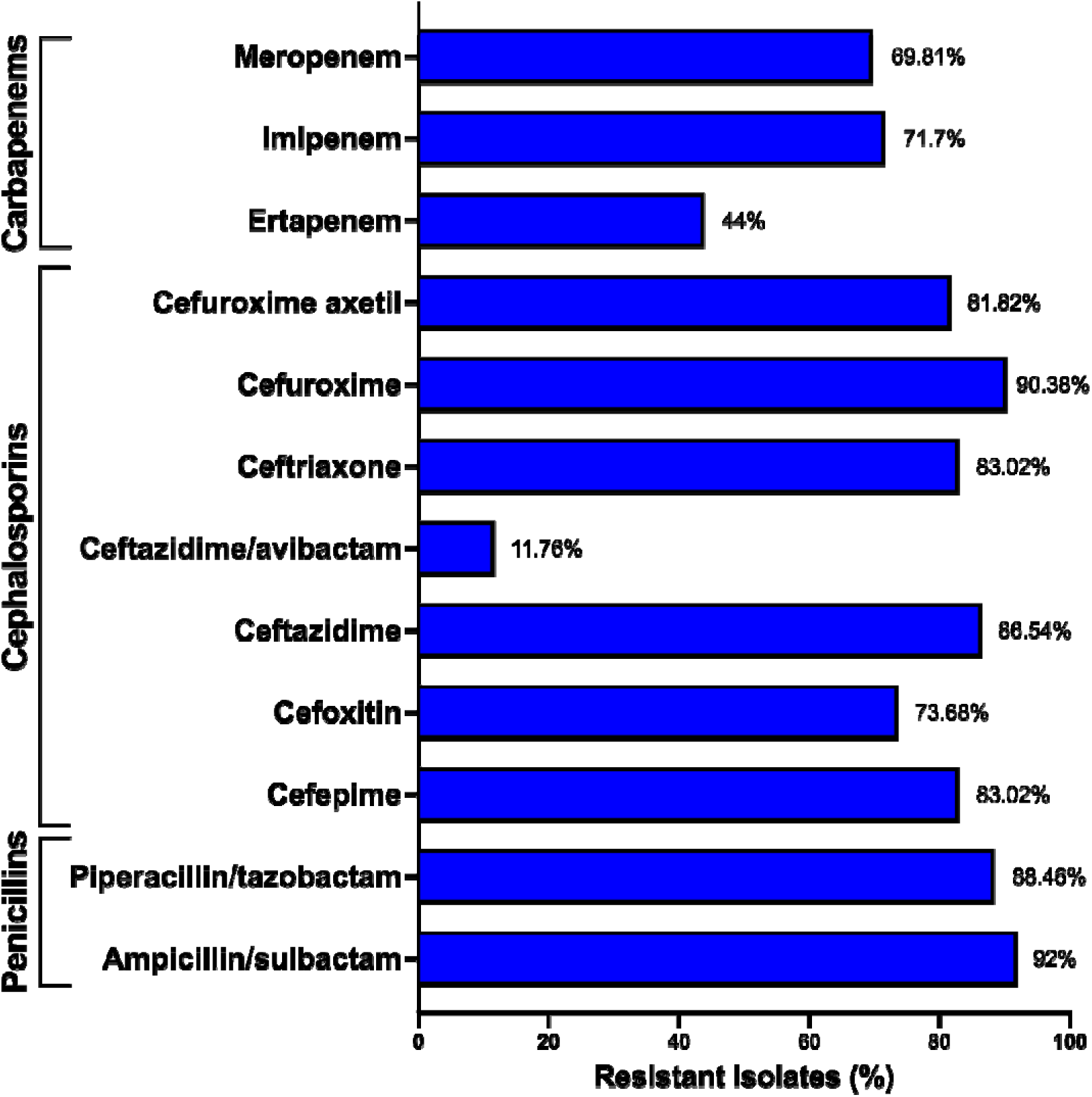
Prevalence of *K. pneumoniae* isolates resistant to the beta-lactams evaluated (Penicillins, Cephalosporins and Carbapenems), demonstrating a phenotypic profile of resistance to multiple beta-lactams drugs, in approximately 70% of the isolates, with the exception of ceftazidime/avibactam and ertapenem.

### Primers design

After selection on the platforms: Comprehensive Antibiotic Resistance Data-base (CARD) and National Center for Biotechnology and Information (NCBI, USA), and subsequent compilation and analysis of FASTA sequences, it was possible to identify consensus sequences common to all sequences available in the databases for each of the genes included in the study: *bla*SHV (N = 156)*, bla*TEM (N = 167)*, bla*NDM (N = 27)*, bla*KPC (N = 208)*, bla*GES (N = 25), and *bla*CTX-M (N = 144). Except for the *blaOXA* gene, for which the most clinically relevant sequences were used *(N = 203)*.

The *bla*CTX-M and *bla*OXA genes, due to their diversity and high degree of genetic variation between homologous sequences, were grouped into clades, and divided into groups. the *bla*CTX-M gene group was named according to the international standardization for this gene with the CTX-M 1*like* been divided into two groups due to its high number or sequences and the genetic variation between them. CTX-M 1.1*like* (N = 42), CTX-M 1.2*like* (N = 16), CTX-M 2*like* (N = 23), CTX-M 8*like* (N = 14) and CTX-M 9*like* (N = 49), while the *blaOXA* gene was divided into the OXA*-23like* (N = 25), OXA*-24/40like* (N = 8), OXA*-48like* (N = 17) and OXA*-51like* (N = 153), as can be seen in **Figures 2** and **3**.

**Figure 2.**
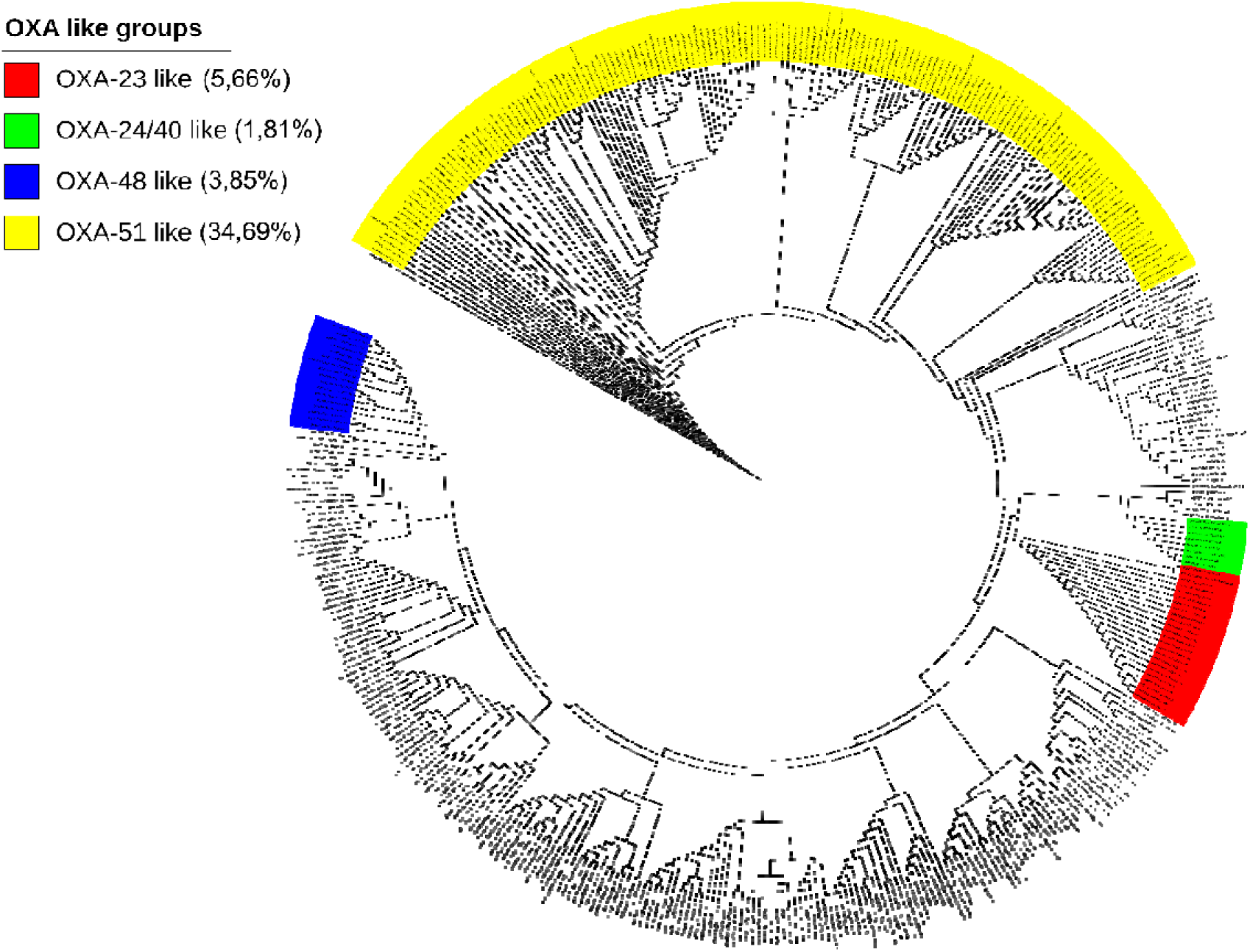
Cladogram of the blaOXA subfamily showing the most epidemiologically relevant groups, covered by the primers developed, and the percentage of sequences identifiable by each primer in comparison with the total number of the blaOXA family (N = 441) at the time of publication of this work.

**Figure 3.**
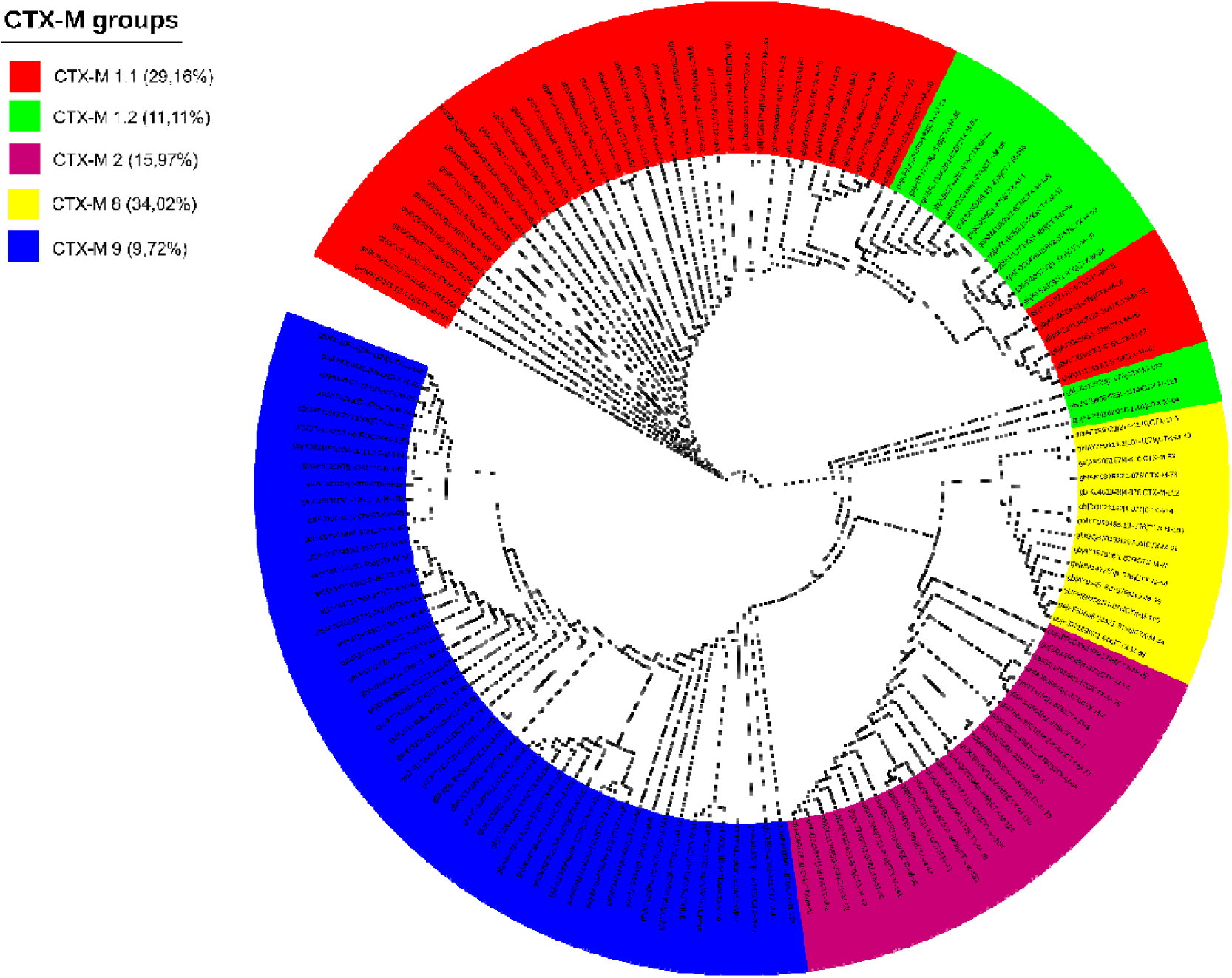
Cladogram with all sequences from the blaCTX-M subfamily divided into groups, illustrating the identifiable sequences, and the percentage of sequences detectable by each primer in comparison with the total number of the blaCTX-M family (N = 144) until the time of publication of this work.

A total of 14 pairs of primers were developed, which together have the capacity to detect 929 variants of resistance genes belonging to the bla gene family. The primer sequences for genes with conserved and non-conserved sequences can be found in Tables 1 and 2, respectively.

**Table 1.**
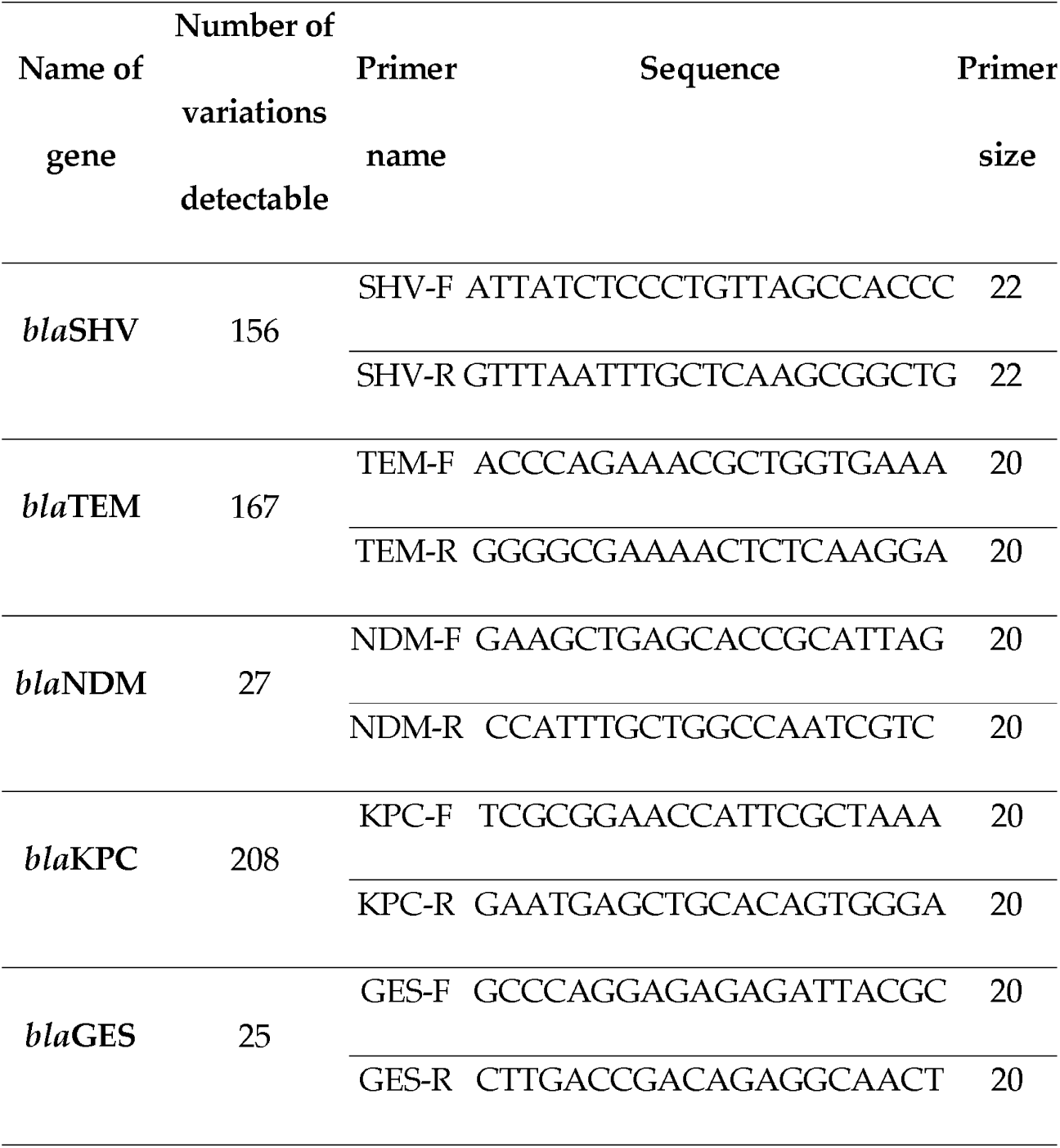
Primer sequences for conserved genes.

**Table 2.**
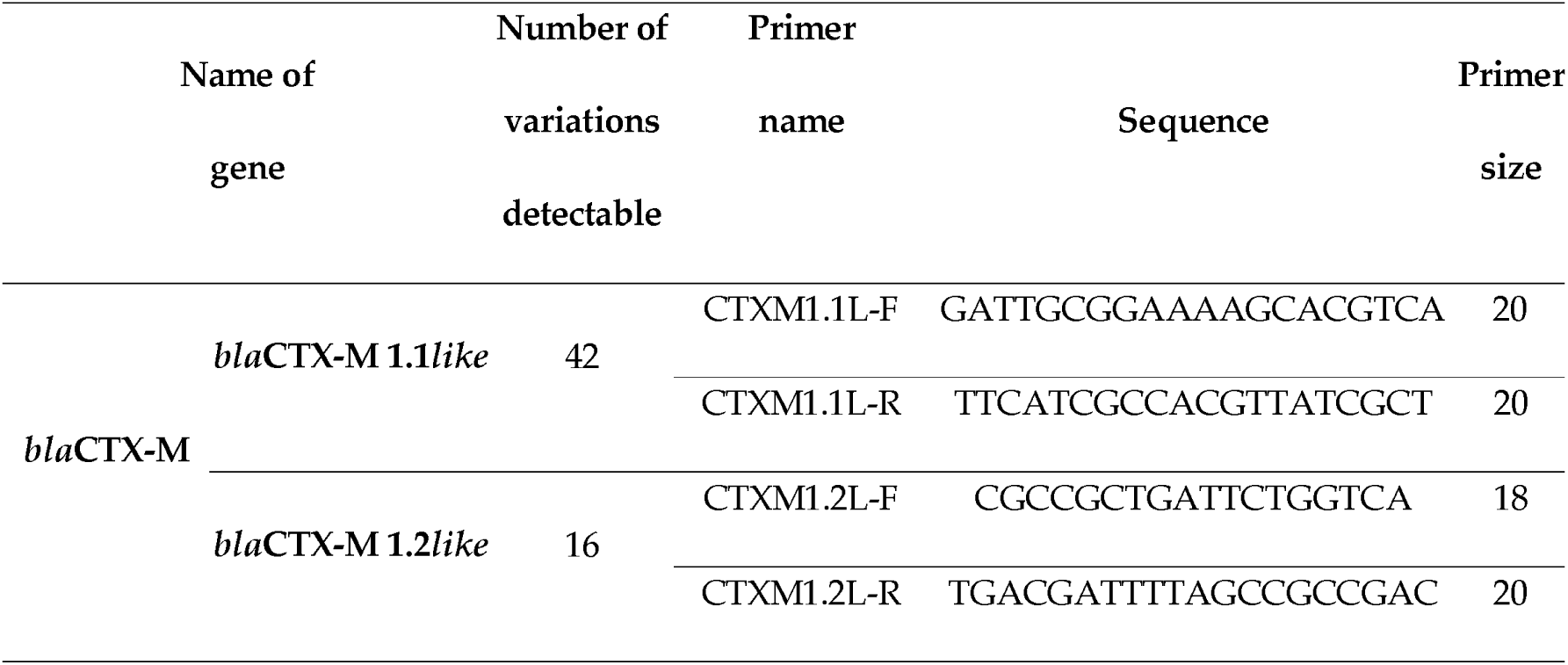
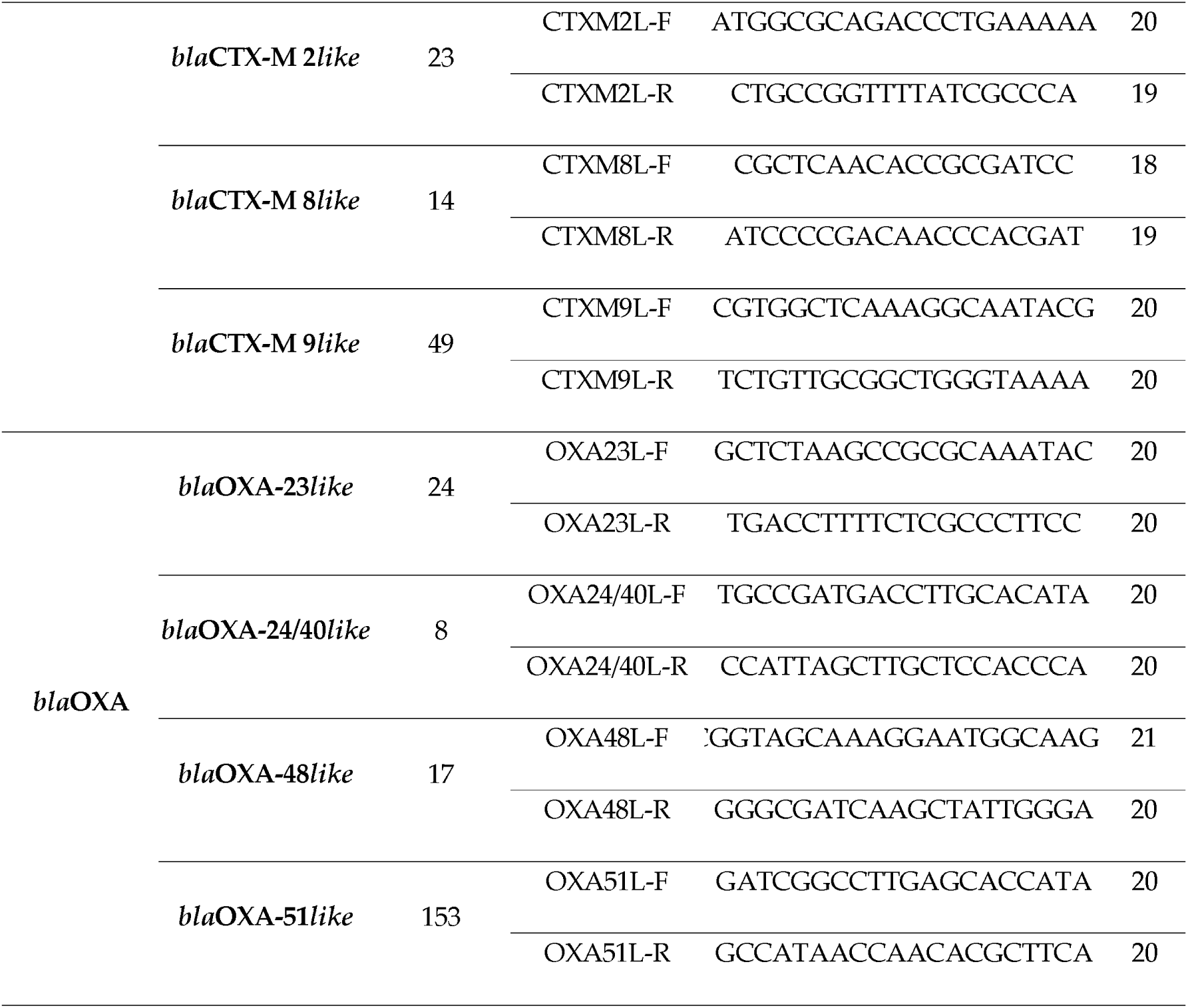
Primer sequences for non-conserved genes, divided into clades.

### *In silico* primers validation

The parameters obtained for each primer after analysis using the Primer-BLAST® (NCBI, USA) and Sequence Manipulation Suite (SMS): PCR Primer Stats software are available in Table 3.

**Table 3.** Parameters obtained by *in silico* validation of the developed primers.

| Gene | Primer name | Tm (C°) | GC% | Self | Self 3' | Product length |
| --- | --- | --- | --- | --- | --- | --- |
|  |  |  |  | complementarity | complementarity |  |
| *bla*SHV | SHV-F | 59,2 | 50,0 | 3 | 0 | 70 |
|  | SHV-R | 59,3 | 45,4 | 5 | 3 |  |
| *bla*TEM | TEM-F | 60,1 | 50,0 | 4 | 0 | 110 |
|  | TEM-R | 59,9 | 55,0 | 2 | 1 |  |
| *bla*NDM | NDM-F | 59,3 | 55,0 | 5 | 1 | 86 |
|  | NDM-R | 60,1 | 55,0 | 6 | 2 |  |
| *bla*KPC | KPC-F | 60,3 | 50,0 | 4 | 2 | 128 |
|  | KPC-R | 60,3 | 55,0 | 5 | 1 |  |
| *bla*GES | GES-F | 59,9 | 60,0 | 3 | 2 | 94 |
|  | GES-R | 59,9 | 55,0 | 3 | 2 |  |
| *bla*CTX-M 1.1*like* | CTXM1.1L-F | 59,7 | 50,0 | 4 | 1 | 87 |
|  | CTXM1.1L-R | 60,1 | 50,0 | 4 | 2 |  |
| *bla*CTX-M 1.2*like* | CTXM1.2L-F | 60,1 | 61,1 | 3 | 2 | 90 |
|  | CTXM1.2L-R | 61,0 | 55,0 | 3 | 3 |  |
| *bla*CTX-M 2*like* | CTXM2L-F | 60,5 | 50,0 | 4 | 0 | 155 |
|  | CTXM2L-R | 60,7 | 57,8 | 4 | 1 |  |
| *bla*CTX-M 8*like* | CTXM8L-F | 61,5 | 66,6 | 4 | 2 | 197 |
|  | CTXM8L-R | 60,9 | 57,8 | 3 | 2 |  |
| *bla*CTX-M 9*like* | CTXM9L-F | 59,9 | 55,0 | 3 | 2 | 180 |
|  | CTXM9L-R | 60,1 | 50,0 | 3 | 0 |  |
| *bla*OXA-23*like* | OXA23L-F | 60,0 | 55,0 | 4 | 0 | 129 |
|  | OXA23L-R | 59,9 | 55,0 | 2 | 0 |  |
| *bla*OXA-24/40*like* | OXA24/40L-F | 59,7 | 50,0 | 4 | 2 | 177 |
|  | OXA24/40L-R | 60,0 | 55,0 | 4 | 0 |  |
| *bla*OXA-48*like* | OXA48L-F | 59,8 | 52,3 | 3 | 0 | 183 |
|  | OXA48L-R | 59,8 | 55,0 | 4 | 0 |  |
| *bla*OXA-51*like* | OXA51L-F | 59,8 | 55,0 | 4 | 2 | 199 |
|  | OXA51L-R | 59,1 | 50,0 | 2 | 1 |  |

### Testing, optimization, and standardization of *in vitro* reactions

All primers were tested using positive controls developed *in house*, and it was possible to verify that the ideal annealing temperature (Ta) for the developed primer pairs was 61°C, except for the primer referring to the *bla*NDM gene, for which the ideal Ta was 64°C. The reactions were then evaluated for their specificity and stability by checking the Melting temperature (Tm). The melting curves are available in the supplementary material **Figure S1**.

### Efficiency curve

It was found that all tested primers presented an efficiency rate ≥93.21 and ≤101.28%, R² ≥0.99, and detection limit between ≈2 and ≈13 copies/µL as shown in **Table 4**.

**Table 4.** Results of the efficiency curves of the developed primers.

| Gene name | Efficiency (%) | Correlation coefficient (R<sup>2</sup>) | Threshold | Detection limit (Copies/μL) | Melting Peak (Tm°C) |
| --- | --- | --- | --- | --- | --- |
| *bla*SHV | 96,15 | 0,992 | 0,60 | $\approx 2$ | 86,7 |
| *bla*TEM | 95,32 | 1,000 | 0,70 | $\approx 2$ | 81,5 |
| *bla*NDM | 99,90 | 0,999 | 0,48 | $\approx 2$ | 85,7 |
| *bla*KPC | 100,44 | 0,992 | 0,24 | $\approx 2$ | 85,7 |
| *bla*GES | 100,21 | 0,997 | 0,27 | $\approx 2$ | 81,7 |
| *bla*CTX-M 1.1like | 95,75 | 0,999 | 0,40 | $\approx 13$ | 84,3 |
| *bla*CTX-M 1.2like | 100,82 | 0,994 | 0,32 | $\approx 13$ | 83,2 |
| *bla*CTX-M 2like | 98,09 | 0,998 | 0,30 | $\approx 2$ | 86,4 |
| *bla*CTX-M 8like | 95,32 | 1,000 | 0,10 | $\approx 13$ | 88,4 |
| *bla*CTX-M 9like | 97,82 | 0,996 | 0,10 | $\approx 2$ | 89,0 |
| *blaOXA-23like* | 99,51 | 1,000 | 0,15 | ≈2 | 78,4 |
| *blaOXA-24/40like* | 101,82 | 0,988 | 0,15 | ≈13 | 78,7 |
| *blaOXA-48like* | 93,21 | 1,000 | 0,20 | ≈13 | 80,29 |
| *blaOXA-51like* | 95,51 | 0,993 | 0,55 | ≈2 | 82,0 |

### Identification of the genetic resistance profile of *K. pneumoniae* isolates

The 122 *K. pneumoniae*isolates were tested against the 14 pairs of primers developed, and it was possible to verify a high prevalence of beta-lactam resistance genes among the isolates analyzed. The most prevalent genes detected were, respectively: *bla*KPC (95.90%); *bla*SHV (94.26%); *bla*CTX-M 1.2*like* (88.52%); *bla*CTX-M 2*like* (83.61%); *bla*CTX-M 1.1*like* (80.33%); *bla*TEM (80.33%); *bla*NDM (33.61%); *bla*OXA-23*like* (28.69%); *bl a*GES (20.49%); *b l a*OXA-51*like* (15.57%); *b l* C*a* TX-M 9*like* (13.93%); *b l a*CTX-M 8*like* (12.30%); *bla*OXA-24/40*like* (11.48%) *and bla*OXA-48*like* (3.28%). The results can be seen in **Figure 4A**, grouped according to Ambler’s classification.

**Figure 4.**
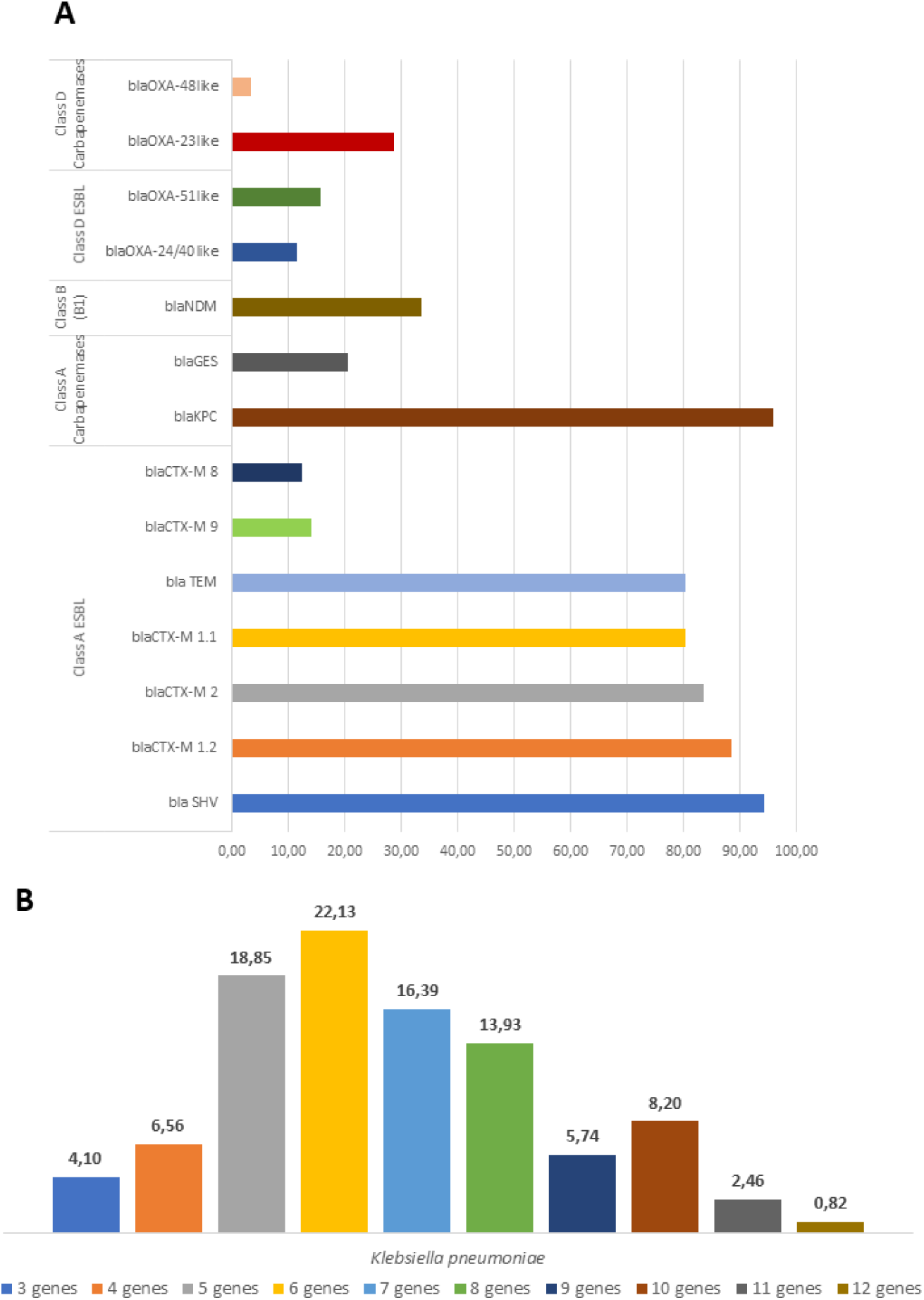
**(A)** Prevalence of beta-lactams resistance genes among the analyzed isolates, grouped according to the Ambler classification. **(B)** The percentage of *K. pneumoniae* isolates showing accumulation of genes encoding beta-lactamases belonging to the *bla* family.

It is important to note that not all variants of each group of genes analyzed belong to a single class in the Ambler’s classification, such as *bla*TEM and *bla*SHV genes, where not all variants have activity as extended spectrum beta-lactamases (ESBL) for example, however, due to the grouping of all sequences in a single primer, the separation of these sequences into other groups would be impossible, so the grouping of genes was carried out globally in a way that covered the mains classification of the subfamily in question.

Through the data obtained by the qPCR reactions developed, it was also possible to verify that the isolates analyzed during this study had the presence of several genes belonging to the *bla* family accumulated, with values varying from 3 to 12 of the tested genes, present per isolate, as shown in **Figure 4B**.

### Comparisons of metabolomic profiles of *K. pneumoniae* isolates

In total, 241 metabolites were identified in positive mode and 116 metabolites identified in negative mode. Among these, 309 metabolites were identified by combining data from both acquisition modes (positive and negative) without redundancy. The principal component analysis (PCAs) and the heatmap analysis between the metabolites in supernatant of susceptible *K. pneumoniae* versus the control culture medium showed complete separation of the main vectors of the PCA plots and differential upregulation or downregulation between the two experimental groups (**Figure 5**). Among these metabolites, 25 were upregulated or downregulated. Interestingly, metabolites related to the synthesis of nitrogenous bases, protein synthesis and energy supply to the bacterial cell are significantly increased in supernatant derived from susceptible *K. pneumoniae* (**Figures S2, S3 and S4**). Similar findings were detected between MDR *K. pneumoniae* strains and control culture medium (**Figures S5, S6 and S7**).

**Figure 5.**
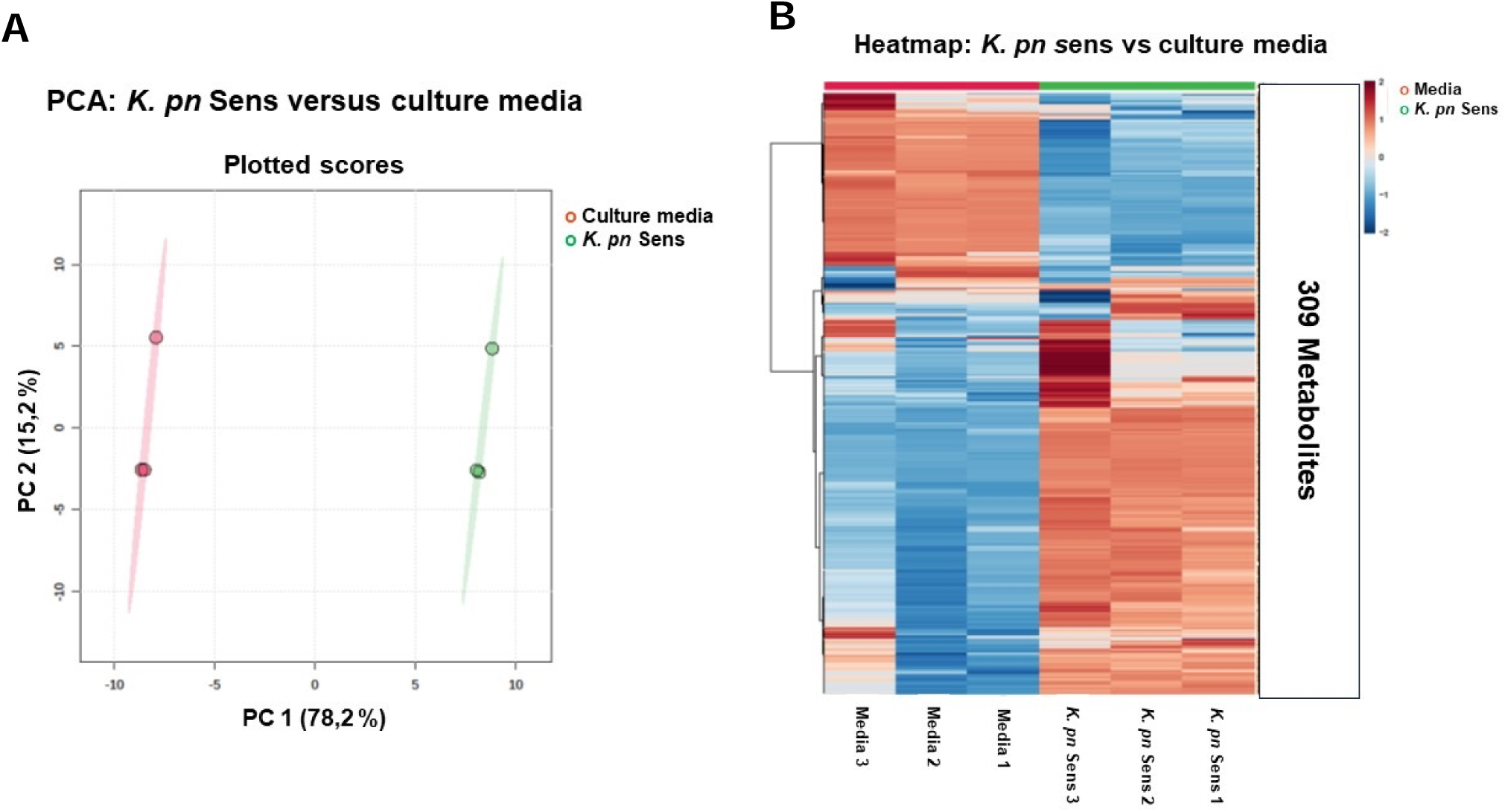
Comparison between susceptible *K. pneumoniae* supernatants (Sens) versus culture media. **(A)** Plot of the principal component analysis (PCAs) and **(B)** the heatmap of the comparison of metabolites between the supernatant of susceptible *K. pneumoniae* versus the control culture media.

The principal component analysis (PCAs) and the heatmap analysis of metabolites in supernatants from *K. pneumoniae* MDR versus susceptible isolates exhibited complete separation of the main vectors of the PCA plots (**Figure 6A**) and differences on levels of metabolites (**Figure 6B**). The analysis of deregulated metabolites between *K. pneumoniae* MDR versus susceptible isolates showed increased N-fructosyl isoleucine in the MDR *K. pneumoniae* isolates (p< 0.05, **Figure 6C**).

**Figure 6.**
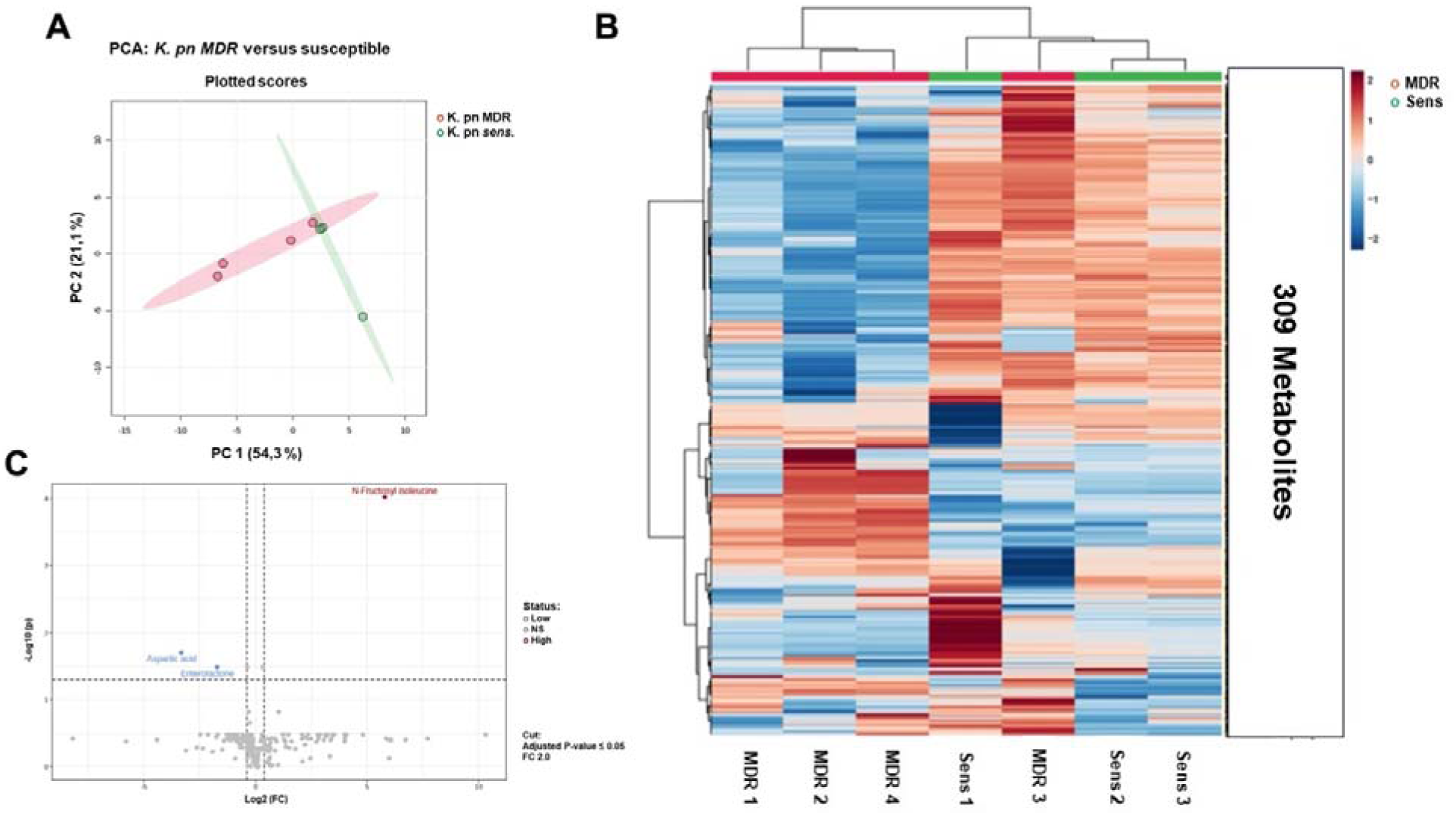
Comparison of supernatants from *K. pneumoniae* MDR versus susceptible (Sens). **(A)** Plots of the principal component analysis (PCAs) and **(B)** the heatmap of the comparison of metabolites between the supernatant of *K. pneumoniae* MDR versus susceptible (Sens). (C) Plots of dysregulated metabolites between supernatants of K. pneumoniae MDR and susceptible.

Analysis of the intracellular metabolites of susceptible versus MDR K. pneumoniae showed differences in the PCAs vectors (Figure 7A), heatmap (Figure 7B). and increased N-fructosyl isoleucine levels in the MDR K. pneumoniae isolates (p<0.05, Figure 7C). Figure 8 shows the PCAs and heatmap plots of supernatant from sensitive and MDR K. pneumoniae.

**Figure 7.**
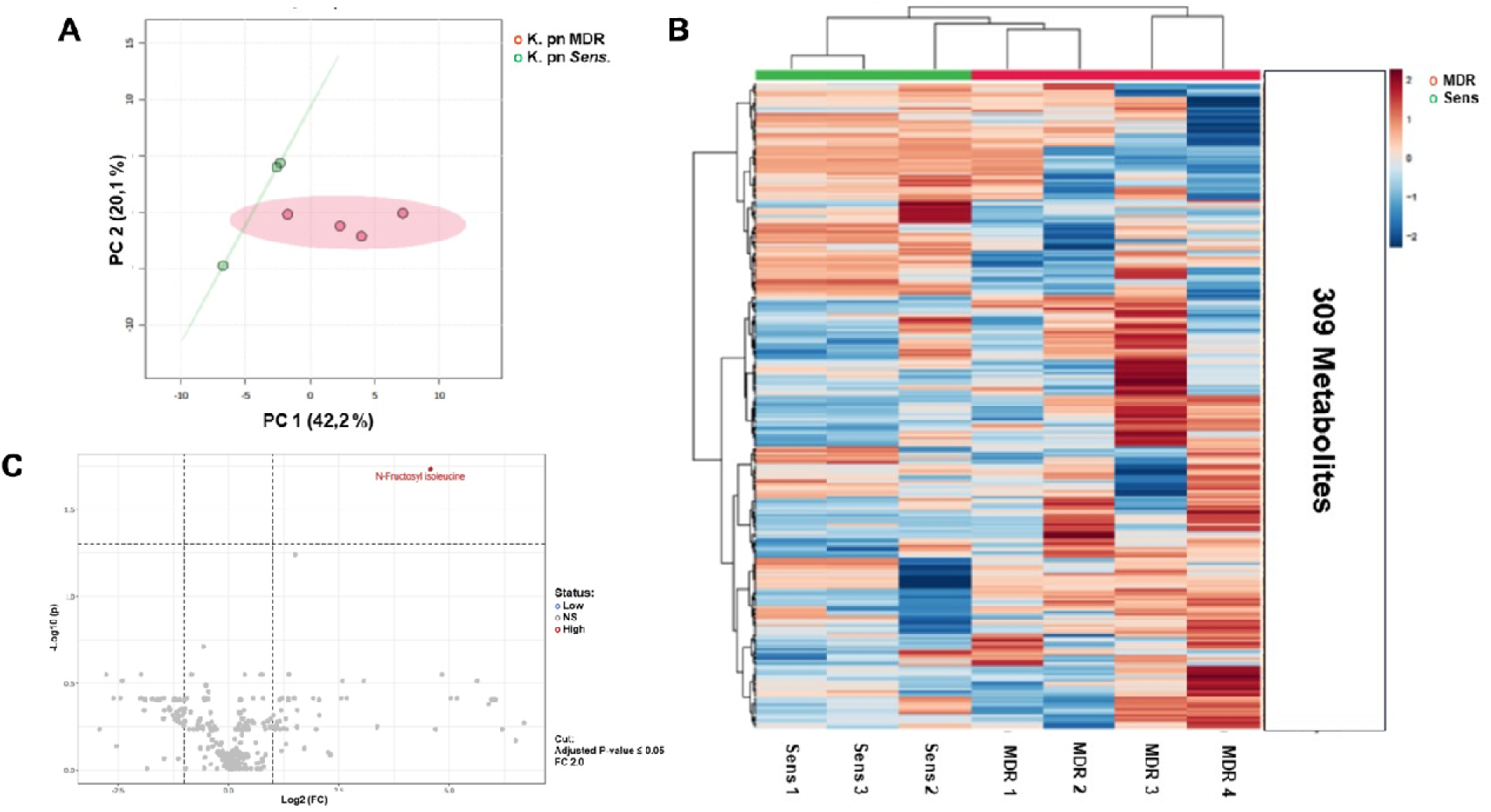
Comparison of Intracellular media of K. pneumoniae MDR versus susceptible (Sens). (A) Principal Component Analysis (PCAs) plots and (B) Heatmap of the Intracellular Environment of K. pneumoniae MDR and susceptible. (C) Plots of dysregulated metabolites between the intracellular media of K. pneumoniae MDR and susceptible.

**Figure 8.**
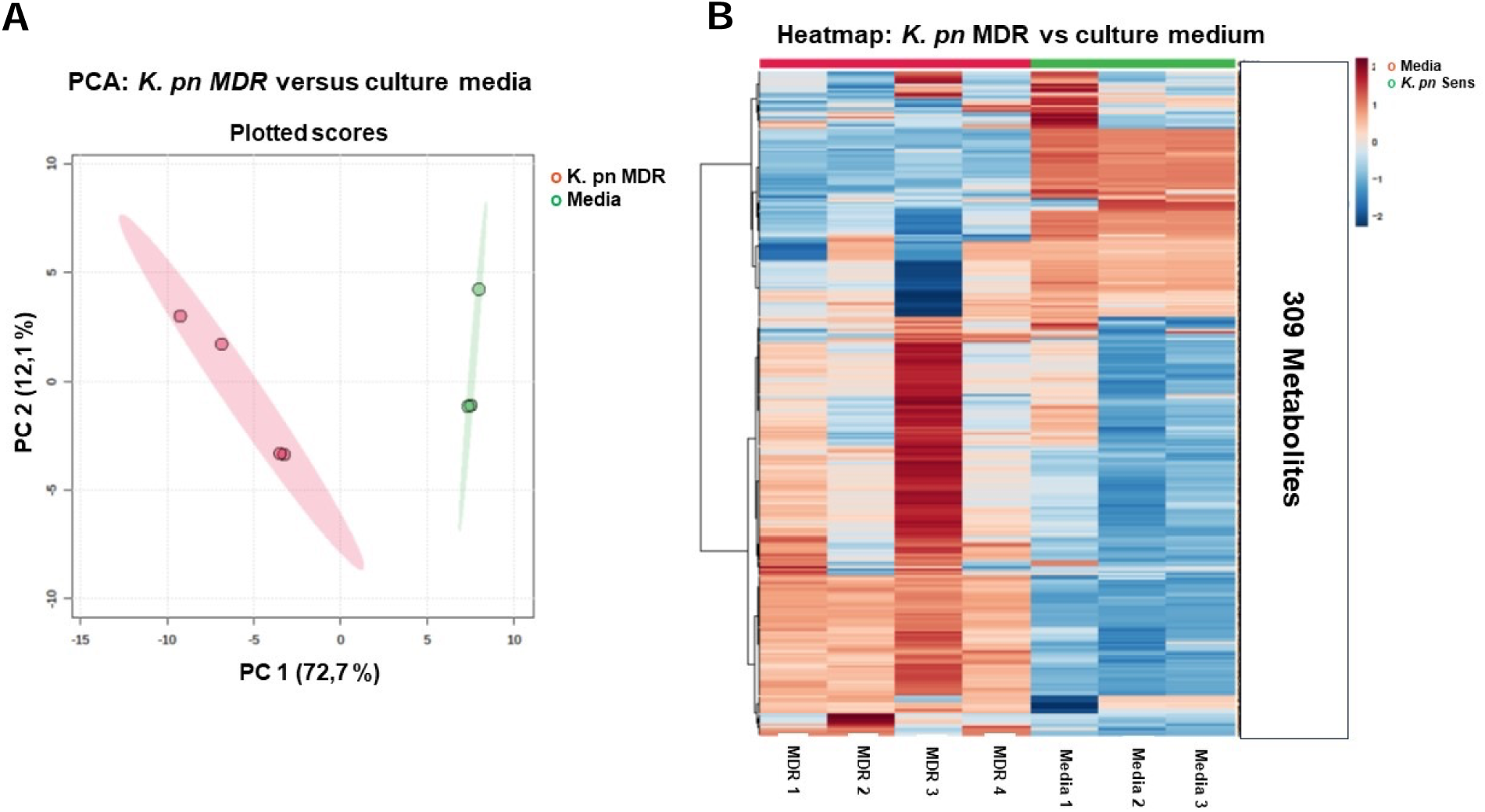
Comparison of *K. pneumoniae* Multidrug-Resistant (MDR) supernatants versus culture media. (A) Principal component analysis (PCAs) plots and (B) Heatmap of the supernatant of *K. pneumoniae* susceptible and MDR. Observe the separation of the main vectors of the (PCAs) plots and details of the differences in the heatmap between the two experimental groups.

## DISCUSSION

During the period analyzed in the study, 249 bacterial isolates resistant to two or more classes of antimicrobials used in TSA were identified. Of these resistant isolates, 48.99% (122/249) belonged to the species *Klebsiella pneumoniae*, being this microorganism the most prevalent among those identified.

This data is consistent with what is seen in Egypt, where when evaluating 186 samples from Cairo hospitals, *K. pneumoniae* was found as the main gram-negative bacterial agent representing 40.9% (76/186) of the identified microorganisms, and among these isolates obtained, a high prevalence of resistance to beta-lactams, quinolones, and sulfonamides was identified (89.4%, 89.4% and 87.1%) respectively.^16^

The phenotypic profile of the *K. pneumoniae* isolates analyzed in the present study demonstrated high levels of resistance to the beta-lactams drugs tested in all classes evaluated (penicillins, cephalosporins and carbapenems). Although the resistance profile of strains may vary depending on the test location and the conditions to prevent spread, the resistance to beta-lactams among *Enterobacteriaceae*is known to be high, as demonstrated in a study carried out in Brazil, with samples of human and veterinary origin where 62.85% and 54.28% of *Enterobacteriaceae*isolated were resistant to amoxicillin/clavulanate and cefazolin respectively.^17^

In studies carried out in Egypt, evaluating isolates of *K. pneumoniae* from food sources, and in Tunisia and Iran evaluating isolates from hospital samples, the presence of high resistance to beta-lactams can be verified for ampicillin/sulbactam (93%), ceftazidime (95.5%), cefoxitin (95.5%), cefotaxime (93.2%), amoxicillin/clavulanate (86.4%), ertapenem (90.9%) and temocillin (84.0%).^18, 19, 20^

Likewise, in a study carried out with 102 isolates obtained from two Portuguese hospitals, the presence of resistance rates > 90% was verified for all beta-lactam drugs tested, except for cephalosporins: cefoxitin (40.2%), cefotetan (68.6%) and carbapenems: ertapenem (23.5%), imipenem (32.4%), meropenem (34.3%) and doripenem (33.3%).^21^

This result is consistent with that found in this study, where the beta-lactam drugs that showed a lower resistance rate belonged to the same classes mentioned, namely: cefoxitin (73.68%), ceftazidime/avibactam (11.76%), ertapenem (44.00%), imipenem (71.70%) and meropenem (69.81%).

Resistance to beta-lactams is conferred in Gram-negatives mainly by genes belonging to the *bla* family. This group of genes is composed of dozens of subfamilies and hundreds of genetic subvariants in each family, the molecular panel developed managed to encompass some of the most predominant and clinically relevant subfamilies among clinical isolates, identified in several studies, namely: *bla*SHV, *bla*TEM, *bla*NDM, *bla*KPC, *bla*GES*, bla*CTX-M and *bla*OXA.^22–24^

After analysis using the Primer-BLAST® (NCBI, USA) and Primer Stats Sequence Manipulation Suite (SMS) software, it was verified that the developed primers have specificity for the sequences used in the alignment, melting temperature (Tm) between 59 29 °C and 61.55 °C, GC% concentration varying between 45.45% and 66.67% in addition to having complementary base numbers ≤ 6 in the analysis of dimer and hairpin formation. The values obtained for these parameters therefore demonstrate that they are compatible with those obtained in reference studies.^25–27^

Due to the high number of sequences included in the development of primers, many of the alternatives obtained through Primer-BLAST® (NCBI, USA) did not meet the stability requirements regarding the formation of secondary structures (self- annealing and hairpins), In order to choose the options with the best performance in relation to these parameters, the GC clamp was relaxed so that the primers SHV-F, SHV-R, CTXM1.2L-R and CTXM2L-R obtained 4 G or C residues within the last 5 nucleotides of the 3’ end, the primers CTXM9L-R and CTXM2L-F did not present C or G residues within the last 5 nucleotides of the sequence.

This fact could lead to the formation of strong bonds and increase the Tm value in primers with more than 3 terminal GC residues, and cause inhibition of amplification due to the presence of weak bonds when there is an absence of terminal CG ^28,29^. Such changes, however, were not evident when the primers were tested *in vitro*.

The variety of sequences detectable by the developed primers makes in vitro validation using positive controls for all sequences virtually impossible. In silico analysis allowed the evaluation of all tested sequences quickly, cheaply and constantly, since the developed primers can be tested against new variants of the target genes.^26–28^

In vitro analyzes were carried out using SYBR Green master mix, the melting curve was therefore used as a parameter to evaluate the specificity of the reactions. And it was verified that for all tested primers a single melting peak was found, indicating the formation of a single amplicon. And excluding the presence of secondary structures (primer dimers and hairpins).

The analysis of the melting curve and Tm is considered fundamental to test the specificity of reactions that use SYBR Green as an amplification indicator, because unlike reactions that use specific probes, which emit fluorescence only when the reaction occurs at the expected target. The SYBR Green works by binding to any double-stranded fragment formed, thus emitting fluorescence. Therefore, only by analyzing the melting curve and the specific Tm of the amplicon is it possible to determine whether the reaction was successful.^29–31^

The efficiency curve and its parameters were used to evaluate the performance of qPCR reactions, by checking how efficiently the targets were amplified in each PCR cycle.^32, 33^ When evaluating the activity of primers in qPCR reactions using the efficiency curve, it was found that all primers analyzed in this study presented efficiency values >90% and <110% and correlation coefficients (R²) >0.9, thus being in accordance with the parameters presented in reference documents present in the literature.^33^ It was also possible to verify a high detection capacity even at low concentrations of genetic material since the detection limit obtained varied between ≈2 and ≈13 copies/µL of the target gene used as control.

Several studies report the growth in the prevalence of the *bla*KPC gene, with this being the most detected carbapenemase worldwide. The values found vary according to the location. The main variant of the *bla*KPC gene detected is *bla*KPC*-2*, with prevalence values varying from 1.2% to 51.6% among enterobacteria in studies carried out in the United States of America and China respectively.^34, 35^ This gene is found more frequently in isolates of *K. pneumoniae*, many studies report high prevalence rates in this species ranging from 17.2% to 64.6%.^4, 35, 36, 37^

### In a study conducted in an intensive care unit in the northeast region of Brazil, it was shown that of 25 isolates of *K.* pneumoniae obtained during the research, 100% exhibited the presence of the *bla*KPC gene. ^38^

The values found in this paper for the *bla*KPC gene may reflect the fact that the primer developed detected all variants of the gene, as well as the higher rates of resistance usually found in intensive care unities (ICU).^39^

The results found in this work are in agreement with other studies performed at the northeast of Brazil^38^, what can represent an endemic presence of the *bla*KPC gene among *K. pneumoniae* isolates in this Brazilian region.

The *bla*TEM and *bla*SHV genes were the first ESBL identified, and together with the *bla*CTX-M subfamily of genes are considered the most prevalent ESBL’s. These genes are widely distributed among enterobacteria; in a study carried out in Sudan, the *bla*TEM, *bla*CTX-M and *bla*SHV genes were identified in 86.0%, 78.0% and 28.0% respectively.^40^

In a work carried out in ICUs in Chile, it was possible to verify the presence of these genes with prevalence rates of 81.0%, 84.7% and 73.0% for *bla*SHV, *bla*CTX-M-1 and *bla*TEM, where a higher prevalence of the *bla*SHV gene was found when compared to the study carried out in Sudan.^41^

In Brazil, it is also possible to verify the presence of high prevalence rates of these genes among *K.* pneumoniae isolates from ICU, with studies indicating rates of 100%, 96% and 72% for *bla*TEM, *bla*SHV and *bla*CTX-M1 respectively. ^38^ These results agree with those evidenced in this study for the K. pneumoniae isolates tested.

The *bla*NDM gene is considered the second most prevalent carbapenemase producing gene in the world, behind only the *bla*KPC gene. In this study, the presence of the metallo beta-lactamase NDM gene was verified in (33.61%) of the isolates. Similar results were evidenced in China, where the presence of this gene was verified in 35.7% of 935 carbapenem-resistant enterobacteria tested.^34^

### The prevalence found in this study for the *bla*NDM gene is still below what was found in other studies carried out in Brazil in the states of São Paulo and Sergipe, where the authors found a prevalence of 70 and 50% respectively in *K. pneumoniae* isolates. ^42, 43^

Regarding oxacillinases, a low prevalence of the *bla*OXA-48*like* gene was observed (3.28%). This gene may be present in all *Enterobacteriaceae*; however, it is more prevalent in strains of *K. pneumoniae*.^21^

The values found in the literature for the presence of this gene in *Enterobacteriaceae* are very divergent, with marked variations related to the location of the analysis, these values vary from 7.3% in China to 83.3% in Morocco.^34, 44^ In a study conducted in Brazil where 4,451 isolates of *Enterobacteriaceae* were analyzed, the *bla*OXA-48*like* gene was detected in only 2.5% of the isolates tested.^45^ In this way, the result obtained is compatible with what was previously identified in Brazil, and close to what is seen in China.

The presence of variants of the *bla*OXA gene (*bla-*OXA-51*like* (15.57%), *bla*OXA- 24/40*like* (11.48%) and *bla*OXA-23*like* (28.69%)) was also observed in the isolates tested, these genes were long considered exclusive to strains of *Acinetobacter baumannii*, however studies have demonstrated the presence of these genes in other *Enterobacteriaceae*, including *K. pneumoniae*.^46^ Results regarding the prevalence of these genes in strains of *K. pneumoniae* are still scarce, however, in a study conducted in Bahrain in the Persian Gulf, the *bla*OXA*-51* and *bla*OXA*-23* genes were identified respectively in 45.8% and 41.6%. % of *K. pneumoniae* isolates analyzed.^47^

It is believed that many of the beta-lactamases that today can be found contained in mobile genetic elements had their origin in chromosomes of other bacteria, as occurred with the SHV type variants that derived from the *K. pneumoniae* chromosomal SHV-1, and the CTX-M type variants that appear to have originated from the chromosomal CTX-M of *Kluyvera* spp^21^. This change in the presentation of these genes, from chromosomal to mobile elements, may explain the appearance of genes such as chromosomal *bla*OXA present in *A. baumannii* strains on plasmids widespread in other species.

It was possible to verify the presence of isolates presenting multiple genes coding for beta-lactamases, from 3 to 12 of the genes analyzed. Studies point to a high prevalence among *Enterobacteriaceae*of the accumulation of genes from the *bla* family, mainly genes belonging to ESBL, OXA beta-lactamases and carbapenemases such as *bla*KPC and *bla*NDM. This phenomenon is more evident in hospital environments where selective pressure favors microorganisms carrying resistance genes and where there is a high rate of sharing of these genes.^48,49^

Investigations into metabolomic modulation in *K. pneumoniae* have demonstrated a promising area for understanding resistance to antimicrobial agents, developing new therapies, and for studying bacterial virulence. For example, significant inhibition of the pentose phosphate pathway, citrate cycle, amino acid and nucleotide metabolism was observed to be beneficial during treatment of MDR *K. pneumoniae* using the bacteriophage-polymyxin combination, showing the potential for new therapeutic targets and pathways metabolic processes to improve the effectiveness of treatment with available antimicrobial agents.^50,51^

This study is in accordance with others that highlighted the importance of the pentose phosphate pathway in MDR *K. pneumoniae* strains.^50^ This finding was present in both media, supernatant and intracellular in MDR isolates, compared with *K. pneumoniae* isolates susceptible to antimicrobial agents. The significant presence of the metabolic N-fructosyl isoleucine (C_12_H_23_NO_7_; MW: 293.316) in the supernatant and intracellular media of *K. pneumoniae* MDR indicates the expression of the fructose degradation enzyme gene (*frwC* ; fructose-specific phosphotransferase system), which is involved in regulating bacterial growth, virulence and overcoming colonization resistance due to the use of alternative carbon source from fructose.^52, 53^

Metabolomics has proven to be a promising approach in the study of antimicrobial resistance. Its use allows for the comprehensive investigation of metabolic changes associates with different resistance mechanisms, such as alternative energy-obtaining pathways, modifications that alter the cell wall, communication between bacterial cells, and changes in the colonization capacity of these microorganisms.^54, 55^

The identification of these metabolic differences plays an important role in the Discovery of new therapeutic targets and strategies for investigating the reversal of resistance in these pathogens.^55–57^ The combination of genetic and metabolomic techniques therefore contributes to a comprehensive view of resistance mechanisms in bacteria. ^56,57^

The presence of N-fructosyl isoleucine in resistant strains, and the metabolic benefit that its presence appears to generate in increasing colonization capacity, together with the selective pressure found in ICU environments due to the constant use of antimicrobials may be related to the high rates of resistant isolates disseminated among patients in this environment.

The metabolic evaluation in turn points to a metabolic advantage in resistant strains compared to sensitive ones, which goes beyond the resistance capacity itself.^58^

This study has some limitations, such as the exploration of only *bla* family genes, limiting a broader understanding of resistance mechanisms to antimicrobial agents. On the other hand, it presents a broad methodology for exploring the important *bla* family genes most associated with resistance to beta-lactams in *K. pneumonia*. The metabolic findings showed a specific indication that suggests an association with virulence and better proteomic studies will be necessary to associate with potential mechanisms of resistance to antimicrobial agents. Furthermore, additional preclinical studies will be necessary to determine and more specifically validate the virulence associated with the pentose phosphate pathway in *K. pneumoniae*.

## CONCLUSIONS

The results of this work show the high prevalence of resistance to antimicrobial agents in isolates of *K. pneumoniae* within the hospital environment, and that this phenotypic pattern exhibited a great concern as it limits the therapeutic options available, directly implicating the patient’s prognosis.

The assays developed were within the quality criteria for qPCR reactions using SYBR Green master mix and were efficient in detecting the beta-lactams resistance genes from the *bla* family evaluated in this study. The genotypes of the *bla* genes obtained are relevant and worrying for the local hospital scenario, and compatible with the identified phenotypic profile.

The identification of the metabolic N-fructosyl isoleucine in the supernatant and intracellular media of *K. pneumoniae* MDR indicates the expression of the fructose degradation enzyme gene, which is involved in regulating bacterial growth, virulence and overcoming resistance to colonization due to the use of an alternative carbon source from fructose.

## MATERIALS AND METHODS

### Obtaining bacterial isolates and identification

For the following study, samples were collected as part of the clinical investigation of patients admitted to the ICU of a tertiary care health unit in the city of Fortaleza-CE. Were included in the work those microorganisms that were identified as gram-negative bacteria, resistant to two or more groups of antimicrobial agents, including subclasses of β-lactams, fluoroquinolones and aminoglycosides.

The bacterial isolates were identified and tested for their susceptibility to antimicrobials using the automated VITEK® 2 Compact method (BioMérieux, Marcy l’Etoile, France), according to the manufacturer’s recommendations. Minimum inhibitory concentrations were interpreted according to the Clinical and Laboratory Standards Institute (CLSI). For quality control of sensitivity tests, strains from the American Type Culture Collection (ATCC) were used. Specimens that had a resistance profile that fit the research objectives were included in the study.

### Extraction of bacterial DNA

To extract the genetic material, the Wizard Genomic DNA Purification extraction and purification kit (Promega, Madison, USA) was used, according to the manufacturer’s recommendations.

After extraction, all samples were then quantified by spectrophotometry using the NanoDropTM 2000 (Thermo Fisher Scientific, Waltham, Massachusetts, USA) and stored in a -80°C freezer until used in the experiments.

### Selection of genes used in the study and obtaining FASTA sequences

For greater coverage of the genetic profile of resistance to β-lactams in gram-negatives, genes with relevant prevalence epidemiology, belonging to the *bla* gene family, were part of the study, and these included the genes*: blaSHV, blaTEM, blaNDM, blaKPC, blaGES, blaCTX-M and blaOXA*. The sequences used were obtained through the Comprehensive Antibiotic Resistance Database (CARD) and National Center for Biotechnology information (NCBI) platforms, which compiles and organizes the resistance gene sequences available in GenBanck, the list of identifiable sequences is available in the supplementary material Text S1.

### Primer design

To design the primers, all variant sequences of each gene included in the study available on the CARD and NCBI platforms were gathered, the sequences were aligned using the Clustal Omega software, and the alignments were analyzed using the SnapGene software.

Consensus regions with homology ≥95% were selected and these were used to design the primers using the Primer Blast platform from the National Center for Biotechnology and Information (NCBI, USA). The consensus sequences obtained is available in the supplementary material Text S2.

Genes with few conserved regions, for which it was not possible to obtain consensus sequences with homology ≥95%, were separated into clades, and primers were then designed for the sequences by phylogenetic grouping, complying with a minimum of 95% similarity.

### *In silico* validation of the developed primers

All primers developed were validated *in silico* regarding their specificity, structure, formation of primer-dimers and hairpins using the Primer-BLAST® (NCBI, USA) and Sequence Manipulation Suite (SMS): PCR Primer Stats platforms.

### Testing, optimization, and standardization of primers

The reactions were standardized with the use of positive controls developed in-house, through amplification of genetic material, isolation, and purification of amplicons, originating from isolates phenotypically resistant to beta-lactams, and negative control (water DNase/RNase free). They were carried out using a CYBR Green master mix (Promega, Madison, USA), the initial results were evaluated regarding the melting temperature (Tm), to confirm the specificity of the amplicons.

To determine the most efficient qPCR conditions, to reduce the existence of non-specificity and facilitate the interpretation of the results, the concentration gradient and annealing temperature (Ta) of the primers were performed. The qPCR reaction conditions included a hot start step at 95°C for 2 minutes, followed by 35 cycles consisting of a denaturation step for 15 seconds at 95°C, and an annealing/extension step for 1 minute at Ta. specific to each primer, all reactions went through a final melting curve step, with a temperature variation of 60 to 95°C with an increase of 0.05°C/sec.

A 9-point efficiency curve was performed with a dilution factor of 1:8, for each primer developed, with concentrations ranging from ≈27,438,596 to ≈2 (copies/µL), through this procedure it was possible to determine the values of threshold, evaluate the efficiency (%), the correlation coefficient (R²) and the limit of detection of each primer.

### Detection of resistance-related genes by molecular biology

The *K. pneumoniae* isolates obtained in the study were tested against the developed primers. Melting curve analysis of all reactions was used to evaluate the specificity of the results of the isolates in comparison to the specific melting temperature (Tm) of the positive control.

### Preparation of *K.* pneumoniae samples for metabolomic evaluation

Isolates of *K. pneumoniae* susceptible and multidrug resistant to antimicrobial agents obtained in this work were submitted for metabolomic analysis. For this experiment, seven samples of the culture supernatant were designed: one sample of the culture medium, two samples of the culture supernatant of susceptible *K. pneumoniae* (biological replicates), four samples of the culture supernatant of multidrug-resistant *K. pneumoniae* (MDR) for untargeted metabolomics determination and analysis. Samples of the intracellular medium of the *K. pneumoniae* culture, two samples of susceptible *K. pneumoniae* (biological replicates) and four samples of MDR *K. pneumoniae* were also used for untargeted metabolomic analysis. Technical replicates were performed for susceptible media and samples to allow for a pilot statistical comparison.

### Methodology for extracting *K. pneumoniae* culture supernatant

To 200 µL of *K. pneumoniae* culture medium or control medium, 800 µL of 80% methanol was added at -20 °C. The samples were vortexed and incubated at -20°C for 2 hours for protein precipitation. The samples were centrifuged for 10 min at 4°C at 10,000 g (Benchtop refrigerated centrifuge 10k RPM Eppendorf) and the supernatants were transferred to new tubes. The samples were dried under vacuum (Speed Vac Thermo fisher) for approximately 4-5 hours. Dried samples were redissolved in 100 µL of 0.1% formic acid in water containing Metabolomics QReSS labeled heavy standards diluted 100X. (https://www.isotope.com/userfiles/files/assetLibrary/MET_RSCH_QReSS.pdf). A quality control (QC) was done, combining 10 µL of each sample. After the QC test in the mass spectrometer, the samples were diluted 10x to be analyzed. The injection volume of each sample was 10 µL per ionization mode.

### Methodology for extractions from the intracellular medium of *K. pneumoniae* culture

To each tube, 750 µL of cold chloroform:methanol (2:1) mixture at -20 °C was added, vortexed and transferred to tubes reinforced with a ball beater. Cells were disrupted in a bead beater with steel balls for 3 min at an intensity of 5 (Bead Ruptor Elite Omni International Material). The tubes were shaken vigorously for 30 min at 4°C in a temperature-controlled thermal shaker (Thermomixer Eppendorf). 400 µL of water was added, shaken vigorously, and centrifuged for 10 min at 10,000 rpm (Benchtop refrigerated centrifuge 10k RPM Eppendorf) for phase separation. The upper aqueous/methanolic phase was saved as a mixture of soluble metabolites and transferred to Eppendorf tubes. Soluble metabolites were dried under vacuum for 3-4 h (Speed Vac Thermo Fisher). Before running, samples were reconstituted in 100 µL of 0.1% formic acid in water containing 100X diluted Metabolomics QReSS labeled heavy standards (https://www.isotope.com/userfiles/files/assetLibrary/MET_RSCH_QReSS.pdf). The QC sample was prepared with 10 µL of each sample. The injection volume of each sample was 10 µL.

### Analysis by Ultra Performance Liquid Chromatography Coupled to Tandem Mass Spectrometry (UPLC-MS/MS) of metabolites

MS data were acquired on the Orbitrap IDX spectrometer (Thermo) connected to the Vanquish UPLC system. Soluble metabolites were separated using a Waters BEH C18 column (100 x 2.1 mm, 1.9 µm) operated at 30 °C and a flow rate of 250 µL/min. Mobile phase A was 0.1% formic acid in water and mobile phase B was 0.1% formic acid in 90% methanol. Bulk scan range: 67-1000 at 120,000 resolutions with 0.6 sec scan range. The 10 most intense ions in each full scan were selected for fragmentation and MS2 spectra were acquired at a resolution of 30,000 and scaled collision dissociation fragmentation energy of 25, 30, 35 was used.

### UPLC gradient

The total time used in the UPLC gradient runs was 15 minutes.

The flow used was 0.250 mL/min and the following gradient in the percentage of mobile phase B was 50% at 8 min, 98% at 9 min. maintaining until a time of 13 min., ending with 0% between a time of 13.1 – 15 min.

### Verification settings in the mass spectrum

The configurations used in the verification through the mass spectrum were orbit trap of the mass spectrum in the master scan, with an orbitrap detector with a resolution of 120,000. We used quadrupole isolation with a scanning range between 67-1000 m/z, using the frequency lens at 60%. The automatic gain control target was customized and normalized to 25%. The maximum injection time mode has been customized with a maximum injection time of 50 ms. MIcroscans of 1 and profile with positive polarity. Font fragmentation has been disabled.

### Data acquisition, metabolic identification, and data analysis

Once the data was acquired, the samples were analyzed using the open-source software MS-DIAL. More details about the software can be obtained here: http://prime.psc.riken.jp/compms/msdial/main.html.^59,60^ Three blank samples were included in the analysis to identify background ions and remove them later. Samples acquired using MS1 full scan used for quantification and data-dependent acquisition (DDA) mode were used for spectral identification of metabolites. Tolerance MS1 was set to 0.01 Da and MS2 set to 0.025 Da. For peak harvesting, the mass slice width was set to 0.1. For peak alignment, the maximum retention time tolerance was set at 0.5 min, the MS1 tolerance was set at 0.01. Peaks were identified by searching MS2 spectra in the public MS-DIAL database downloaded in January 2023 (324,191 records for positive mode and 64,669 entries for negative mode) using a mass tolerance of 0.01 Da for MS1 and 0.05 Gives to MSMS with an identification cutoff point of 80% http://prime.psc.riken.jp/compms/msdial/main.html#MSP. Additionally, peaks were also searched against the core Biomolecular Analysis Facility’s internal IROA library (both positive and negative mode) with a mass tolerance of 0.02 and identification

cutoff of 85%. The data was manually inspected and identifications without MS2 were filtered, except for identifications of the IROA molecule.

## Supplementary Materials

The following supporting information: Figures S1-S7; Text S1; Text S2.

## Author Contributions

- <u>Bibliographic review and data collection</u>, Lavouisier F.B. Nogueira, Marília S. Maia, Marco A.F Clementino and Aldo A.M. Lima.
- <u>Methodology</u>, Lavouisier F.B. Nogueira, Marília S. Maia, Marco A.F Clementino, Alexandre Havt, Ila F.N. Lima, Jorge L.N. Rodrigues, Luciana V. C. Fragoso and Aldo A. M. Lima.
- <u>Bioinformatics</u>, Lavouisier F.B. Nogueira and Marco A.F Clementino.
- <u>In silico analyses</u>, Lavouisier F.B. Nogueira, Marco A.F Clementino.
- *In vitro* validation of primers and sample testing, Lavouisier F.B. Nogueira, Marília S. Maia.
- <u>Metabolomics, Datascience and data Bank</u>, Jose Q.S. Filho; Deiziane V.S. Costa, José K. Sousa, Lyvia M.V.C. Magalhães, Dilza Silva, Nicholas E. Sherman and Aldo A.M. Lima
- <u>Written review and editing</u>, Lavouisier F.B. Nogueira.
- <u>Supervision</u>, Alexandre Havt and Aldo A.M. Lima.
- Project administration, Aldo A.M. Lima.
- <u>Resource acquisition</u>, Aldo A.M. Lima.

## Acknowledge

This work used the following equipment’s (Benchtop refrigerated centrifuge 10k RPM Eppendorf; Speed Vac Thermo fisher; Bead Ruptor Elite Omni International Material; Thermomixer Eppendorf; Orbitrap IDX spectrometer (Thermo) connected to the Vanquish UPLC system) available in the Biomolecular Analysis Core Facility which is supported by the University of Virginia School of Medicine, Research Resource Identifiers (RRID):SCR_025476.

## Data Availability Statement

The data and reports relating to this study will be available via direct request via email to the corresponding author.

