## Supplemental material for "Molecular Insights into β-Lactams Resistance in *Klebsiella pneumoniae* Clinical Isolates with a Focus on Multidrug Resistance and Virulence"

**Table of contents:**

**1)** Supplementary **Text S1**: Identifiable sequences for each gene included in the study available on the CARD and NCBI platforms.	**pg 3-12**

**2)** Supplementary **Text S2**: Consensus sequences obtained for the gene groups analyzed.	**pg 13-17**

**3)** Supplementary **Figure S1**: Primer’s specificity evaluation through Melting curve: (A) SHV, (B) KPC, (C) NDM, (D) TEM, (E) GES, (F) OXA-23, (G) OXA-24/40, (H) OXA-48, (I) OXA-51, (J) CTX-M 1.1*like*, (K) CTX-M 1.2*like*, (L) CTX-M 2*like*, (M) CTX-M 8*like* e (N) CTX-M 9*like*.	**pg 18**

**4)** Supplementary **Figure S2**: Plot analysis of the 25 main metabolites from the supernatant of susceptible K. pneumoniae in general without separation by up- or down-regulation of the analytes. Note that in the supernatant of *K. pneumoniae* compared to the culture media, metabolites related to the synthesis of nitrogenous bases, protein synthesis and energy supply to the bacterial cell are significantly increased.	**pg 19**

**5)** Supplementary **Figure S3**: Plot of the analysis of the 25 main metabolites from the supernatant of susceptible *K. pneumoniae,* separated because they were up-regulated in relation to the control culture media.	**pg 20**

**6)** Supplementary **Figure S4**: Plot of the analysis of the 25 main metabolites from the supernatant of susceptible *K. pneumoniae,* separated because they were down-regulated in relation to the control culture media.	**pg 21**

**7)** Supplementary **Figure S5**: Plot of the analysis of the 25 main metabolites from the supernatant of Multidrug-Resistant (MDR) *K. pneumoniae,* in general without separation by up- or down-regulation of the analytes. Note that in the intracellular media of K. pneumoniae compared to the culture medium, metabolites related to the synthesis of nitrogenous bases, protein synthesis and energy supply to the bacterial cell are significantly increased.	**pg 22**

**8)** Supplementary **Figure S6**: Plot of the analysis of the 25 main metabolites from the supernatant of Multidrug-Resistant (MDR) *K. pneumoniae,* separated by being up-regulated in relation to the control culture media.	**pg 23**

**9)** Supplementary **Figure S7**: Plot of the analysis of the 25 main metabolites from the supernatant of *K. pneumoniae* Multidrug-Resistant (MDR)*,* separated because they were down-regulated in relation to the control culture media.	**pg 24**

**Text S1:** Identifiable sequences for each gene included in the study available on the CARD platform.

***bla*SHV Gene:**

>gb|FJ668814|77-937|SHV-1 [Klebsiella pneumoniae]; >gb|AF148851|6-866|SHV-2 [Escherichia coli]; >gb|X98102|74-934|SHV-2A [Klebsiella pneumoniae]; >gb|KX092356.1|193-1053|SHV-3 [Klebsiella pneumoniae]; >gb|LT985229.1|36108-36968|SHV-4 [Escherichia coli]; >gb|X55640|112-972|SHV-5 [Klebsiella pneumoniae]; >gb|Y11069|1-780|SHV-6 [Klebsiella pneumoniae]; >gb|U20270|125-985|SHV-7 [Escherichia coli]; >gb|U92041|1-861|SHV-8 [Escherichia coli]; >gb|S82452|121-978|SHV-9 [Klebsiella pneumoniae]; >gb|X98101|74-934|SHV-11 [Klebsiella pneumoniae]; >gb|AJ920369|24-860|SHV-12 [Escherichia coli]; >gb|AF164577|1-861|SHV-13 [Klebsiella pneumoniae]; >gb|AF226622|55-915|SHV-14 [Klebsiella pneumoniae]; >gb|AJ011428|1-861|SHV-15 [Escherichia coli]; >gb|AF072684|118-993|SHV-16 [Klebsiella pneumoniae]; >gb|AF132290|88-948|SHV-18 [Klebsiella pneumoniae]; >gb|AF117743|1-780|SHV-19 [Klebsiella pneumoniae]; >gb|AF117744|1-780|SHV-20 [Klebsiella pneumoniae]; >gb|AF117745|1-780|SHV-21 [Klebsiella pneumoniae]; >gb|AF117746|1-780|SHV-22 [Klebsiella pneumoniae]; >gb|AF117747|1-780|SHV-23 [Klebsiella pneumoniae]; >gb|AB023477|1-861|SHV-24 [Escherichia coli]; >gb|AF208796|1-861|SHV-25 [Klebsiella pneumoniae]; >gb|AF227204|74-934|SHV-26 [Klebsiella pneumoniae]; >gb|AF293345|1-861|SHV-27 [Klebsiella pneumoniae]; >gb|AF299299|1-861|SHV-28 [Klebsiella pneumoniae]; >gb|AF301532|8-868|SHV-29 [Klebsiella pneumoniae]; >gb|AY661885|49-909|SHV-30 [Enterobacter cloacae]; >gb|AY277255|67-927|SHV-31 [Klebsiella pneumoniae]; >gb|AY037778|92-952|SHV-32 [Klebsiella pneumoniae]; >gb|JX268631|1-861|SHV-33 [Klebsiella pneumoniae]; >gb|AY036620|89-949|SHV-34 [Escherichia coli]; >gb|AY070258|1-861|SHV-35 [Klebsiella pneumoniae]; >gb|AF467947|1-861|SHV-36 [Klebsiella pneumoniae]; >gb|AF467948|1-861|SHV-37 [Klebsiella pneumoniae]; >gb|AY079099|149-1009|SHV-38 [Klebsiella pneumoniae]; >gb|AF535128|1-861|SHV-40 [Klebsiella pneumoniae]; >gb|AF535129|1-861|SHV-41 [Klebsiella pneumoniae]; >gb|AF535130|1-861|SHV-42 [Klebsiella pneumoniae]; >gb|AY065991|7-867|SHV-43 [Klebsiella pneumoniae]; >gb|AY259119|117-977|SHV-44 [Klebsiella pneumoniae]; >gb|AF547625|1-861|SHV-45 [Klebsiella pneumoniae]; >gb|AY210887|112-972|SHV-46 [Klebsiella oxytoca]; >gb|AY263404|1-861|SHV-48 [Klebsiella pneumoniae]; >gb|AY528718|414-1274|SHV-49 [Klebsiella pneumoniae]; >gb|AY288915|1-861|SHV-50 [Klebsiella pneumoniae]; >gb|AY289548|1-861|SHV-51 [Klebsiella pneumoniae]; >gb|HQ845196|1-861|SHV-52 [Klebsiella pneumoniae]; >gb|AY590467|1-729|SHV-53 [Klebsiella pneumoniae]; >gb|AJ863560|1-861|SHV-55 [Klebsiella pneumoniae]; >gb|EU586041|1-861|SHV-56 [Klebsiella pneumoniae]; >gb|AY223863|171-1031|SHV-57 [Escherichia coli]; >gb|AY790341|1-861|SHV-59 [Klebsiella pneumoniae]; >gb|AB302939|9-869|SHV-60 [Klebsiella pneumoniae]; >gb|AJ866284|1-861|SHV-61 [Klebsiella pneumoniae]; >gb|AJ866285|1-861|SHV-62 [Klebsiella pneumoniae]; >gb|EU342351|173-1033|SHV-63 [Klebsiella pneumoniae]; >gb|DQ174304|5-865|SHV-64 [Klebsiella pneumoniae]; >gb|DQ174305|5-865|SHV-65 [Klebsiella pneumoniae]; >gb|DQ174306|5-865|SHV-66 [Klebsiella pneumoniae]; >gb|DQ174307|5-865|SHV-67 [Klebsiella pneumoniae]; >gb|DQ174308|5-865|SHV-69 [Klebsiella pneumoniae]; >gb|DQ013287|1-861|SHV-70 [Enterobacter cloacae]; >gb|AM176546|1-861|SHV-71 [Klebsiella pneumoniae]; >gb|AM176547|31-891|SHV-72 [Klebsiella pneumoniae]; >gb|AM176548|17-874|SHV-73 [Klebsiella pneumoniae]; >gb|AM176549|31-891|SHV-74 [Klebsiella pneumoniae]; >gb|AM176550|31-891|SHV-75 [Klebsiella pneumoniae]; >gb|AM176551|31-891|SHV-76 [Klebsiella pneumoniae]; >gb|AM176552|31-891|SHV-77 [Klebsiella pneumoniae]; >gb|AM176553|31-891|SHV-78 [Klebsiella pneumoniae]; >gb|AM176554|31-891|SHV-79 [Klebsiella pneumoniae]; >gb|AM176555|1-861|SHV-80 [Klebsiella pneumoniae]; >gb|AM176556|26-886|SHV-81 [Klebsiella pneumoniae]; >gb|AM176557|1-861|SHV-82 [Klebsiella pneumoniae]; >gb|AM176558|1-861|SHV-83 [Klebsiella pneumoniae]; >gb|AM087453|26-856|SHV-84 [Escherichia coli]; >gb|DQ322460|16-876|SHV-85 [Klebsiella pneumoniae]; >gb|DQ328802|1-861|SHV-86 [Klebsiella pneumoniae]; >gb|DQ193536|1-861|SHV-89 [Klebsiella pneumoniae]; >gb|DQ836922|1-861|SHV-92 [Klebsiella pneumoniae]; >gb|EF373969|1-861|SHV-93 [Klebsiella pneumoniae]; >gb|EF373970|1-861|SHV-94 [Klebsiella pneumoniae]; >gb|EF373972|1-861|SHV-95 [Citrobacter freundii]; >gb|EF373971|1-861|SHV-96 [Acinetobacter baumannii]; >gb|EF373973|1-861|SHV-97 [Enterococcus faecalis]; >gb|EU155018|1-861|SHV-101 [Klebsiella pneumoniae]; >gb|EU024485|1-861|SHV-102 [Escherichia coli]; >gb|EU032604|1-861|SHV-103 [Klebsiella pneumoniae]; >gb|EU274581|1-861|SHV-104 [Klebsiella pneumoniae]; >gb|FJ194944|47-907|SHV-105 [Klebsiella pneumoniae]; >gb|AM941847|1-861|SHV-106 [Klebsiella pneumoniae]; >gb|AM941848|1-861|SHV-107 [Klebsiella pneumoniae]; >gb|HM751100|1-861|SHV-108 [Klebsiella pneumoniae]; >gb|EU418913|17-877|SHV-109 [Klebsiella pneumoniae]; >gb|HQ877615|1-861|SHV-110 [Klebsiella pneumoniae]; >gb|AB372881|9-869|SHV-111 [Klebsiella pneumoniae]; >gb|JF812965|1-861|SHV-120 [Escherichia coli]; >gb|HQ661362|72-932|SHV-121 [Klebsiella pneumoniae]; >gb|HM751103.1|1-864|SHV-122 [Klebsiella pneumoniae]; >gb|GQ390805|1-813|SHV-123 [Klebsiella pneumoniae]; >gb|GQ390806|1-813|SHV-124 [Klebsiella pneumoniae]; >gb|GQ390807|1-813|SHV-125 [Klebsiella pneumoniae]; >gb|GQ390808|1-813|SHV-126 [Escherichia coli]; >gb|GQ390809|1-813|SHV-127 [Klebsiella pneumoniae]; >gb|GU932590|1-861|SHV-128 [Enterobacter cloacae]; >gb|GU827715|1-861|SHV-129 [Escherichia coli]; >gb|AB551737|15-875|SHV-133 [Klebsiella pneumoniae]; >gb|HM559945|1-861|SHV-134 [Klebsiella pneumoniae]; >gb|HQ637576|1-861|SHV-135 [Escherichia coli]; >gb|HQ661363|72-932|SHV-137 [Klebsiella pneumoniae]; >gb|JN051143|1-861|SHV-140 [Klebsiella pneumoniae]; >gb|JQ388884|1-861|SHV-141 [Klebsiella pneumoniae]; >gb|JQ029959|29-889|SHV-142 [Klebsiella pneumoniae]; >gb|JX013655|1-861|SHV-145 [Klebsiella pneumoniae]; >gb|JX121114|1-858|SHV-147 [Klebsiella pneumoniae]; >gb|JX121115|1-858|SHV-148 [Klebsiella pneumoniae]; >gb|JX121116|1-858|SHV-149 [Klebsiella pneumoniae]; >gb|JX121117|1-858|SHV-150 [Klebsiella pneumoniae]; >gb|JX121118|1-858|SHV-151 [Klebsiella pneumoniae]; >gb|JX121119|1-858|SHV-152 [Klebsiella pneumoniae]; >gb|JX121120|1-858|SHV-153 [Klebsiella pneumoniae]; >gb|JX121121|1-858|SHV-154 [Klebsiella pneumoniae]; >gb|JX121122|1-858|SHV-155 [Klebsiella pneumoniae]; >gb|JX121123|1-861|SHV-156 [Klebsiella pneumoniae]; >gb|JX121124|1-861|SHV-157 [Klebsiella pneumoniae]; >gb|JX121125|1-858|SHV-158 [Klebsiella pneumoniae]; >gb|JX121126|1-858|SHV-159 [Klebsiella pneumoniae]; >gb|JX121127|1-861|SHV-160 [Klebsiella pneumoniae]; >gb|JX121128|1-861|SHV-161 [Klebsiella pneumoniae]; >gb|JX121129|1-858|SHV-162 [Klebsiella pneumoniae]; >gb|JX121130|1-858|SHV-163 [Klebsiella pneumoniae]; >gb|JX121131|1-858|SHV-165 [Klebsiella pneumoniae]; >gb|AB733453|4-849|SHV-167 [Klebsiella pneumoniae]; >gb|AM941844|1-861|SHV-98 [Klebsiella pneumoniae]; >gb|AM941845|1-861|SHV-99 [Klebsiella pneumoniae]; >gb|AM941846|1-900|SHV-100 [Klebsiella pneumoniae]; >gb|KJ776406|17-877|SHV-119 [Klebsiella pneumoniae]; >gb|JQ341060.1|882-1742|SHV-143 [Klebsiella pneumoniae]; >gb|JQ926986|1-861|SHV-144 [Klebsiella pneumoniae]; >gb|HE981194|1-861|SHV-164 [Klebsiella pneumoniae]; >gb|KP050487.1|1-861|SHV-180 [Klebsiella pneumoniae]; >gb|HG934764|1-864|SHV-183 [Enterobacter cloacae]; >gb|JX870080|1-861|SHV-168 [Klebsiella pneumoniae]; >gb|KF513177|1-861|SHV-172 [Klebsiella pneumoniae]; >gb|KF513178|1-861|SHV-173 [Klebsiella pneumoniae]; >gb|KF705209|1-861|SHV-178 [Klebsiella pneumoniae]; >gb|KF705208|1-861|SHV-179 [Klebsiella pneumoniae]; >gb|KP050489|1-861|SHV-182 [Klebsiella pneumoniae]; >gb|KM233164|1-861|SHV-185 [Klebsiella pneumoniae]; >gb|KM233165|1-861|SHV-186 [Klebsiella pneumoniae]; >gb|LN515533|1-867|SHV-187 [Klebsiella pneumoniae]; >gb|LN515534|1-873|SHV-188 [Klebsiella pneumoniae]; >gb|KP050494|1-861|SHV-189 [Klebsiella pneumoniae].

***bla*TEM Gene:**

>gb|AL513383|161911-162771|TEM-1 [Salmonella enterica subsp. enterica serovar Typhi str. CT18]; >gb|X54606|215-1075|TEM-2 [Pseudomonas aeruginosa]; >gb|X64523.1|477-1337|TEM-3 [Klebsiella pneumoniae]; >gb|LK391770.1|19708-20568|TEM-4 [Klebsiella pneumoniae]; >gb|X57972|340-1200|TEM-6 [Escherichia coli]; >gb|AF527798.1|1-785|TEM-7 [Escherichia coli]; >gb|X65252|176-1036|TEM-8 [Klebsiella pneumoniae]; >gb|AF093512|198-1058|TEM-10 [Morganella morganii]; >gb|AY874537|178-1038|TEM-11 [Proteus mirabilis]; >gb|M88143|368-1228|TEM-12 [Klebsiella oxytoca]; >gb|AM849805|263-1123|TEM-15 [Haemophilus parainfluenzae]; >gb|X65254|176-1036|TEM-16 [Klebsiella pneumoniae]; >gb|Y14574|1-861|TEM-17 [Capnocytophaga ochracea]; >gb|JX042489|142-1002|TEM-19 [Acinetobacter baumannii]; >gb|Y17581|79-936|TEM-20 [Klebsiella pneumoniae]; >gb|Y17582|1-858|TEM-21 [Klebsiella pneumoniae]; >gb|Y17583|214-1071|TEM-22 [Klebsiella pneumoniae]; >gb|X65253|176-1036|TEM-24 [Klebsiella pneumoniae]; >gb|NG_050256.1|101-961|TEM-26 [Enterobacteriaceae]; >gb|U37195|76-936|TEM-28 [Escherichia coli]; >gb|Y17584|1-858|TEM-29 [Escherichia coli]; >gb|AJ437107|209-1069|TEM-30 [Escherichia coli]; >gb|GU371926|50305-51165|TEM-33 [Escherichia coli]; >gb|KC292503|4335-5195|TEM-34 [Haemophilus parainfluenzae]; >gb|FR717535|1-861|TEM-40 [Escherichia coli]; >gb|X98047|1-844|TEM-42 [Pseudomonas aeruginosa]; >gb|U95363|1-861|TEM-43 [Klebsiella pneumoniae]; >gb|X95401|209-1069|TEM-45 [Escherichia coli]; >gb|Y10279|1-861|TEM-47 [Klebsiella pneumoniae]; >gb|Y10280|1-861|TEM-48 [Klebsiella pneumoniae]; >gb|Y10281|1-861|TEM-49 [Escherichia coli]; >gb|Y13612|1-861|TEM-52 [Klebsiella pneumoniae]; >gb|AF104441|194-1054|TEM-53 [Klebsiella pneumoniae]; >gb|AF104442|194-1054|TEM-54 [Escherichia coli]; >gb|DQ286729|1-861|TEM-55 [Escherichia coli]; >gb|FJ405211|1-861|TEM-57 [Escherichia coli]; >gb|AF062386|31-862|TEM-59 [Klebsiella oxytoca]; >gb|AF047171|136-996|TEM-60 [Providencia stuartii]; >gb|AF332513|104-964|TEM-63 [Escherichia coli]; >gb|AF091113|451-1311|TEM-67 [Proteus mirabilis]; >gb|AJ239002|1-861|TEM-68 [Klebsiella pneumoniae]; >gb|AF188199|215-1075|TEM-70 [Escherichia coli]; >gb|AF203816|211-1071|TEM-71 [Klebsiella pneumoniae]; >gb|AF157553|148-1008|TEM-72 [Morganella morganii]; >gb|AJ012256|209-1069|TEM-73 [Proteus mirabilis]; >gb|AY130284|1-785|TEM-75 [Klebsiella pneumoniae]; >gb|AF190694|209-1069|TEM-76 [Escherichia coli]; >gb|AF190693|209-1069|TEM-78 [Escherichia coli]; >gb|AF190692|209-1069|TEM-79 [Escherichia coli]; >gb|AF347054|209-1069|TEM-80 [Enterobacter cloacae]; >gb|AF427127|209-1069|TEM-81 [Escherichia coli]; >gb|AF427128|209-1069|TEM-82 [Escherichia coli]; >gb|AF427129|209-1069|TEM-83 [Escherichia coli]; >gb|AF427130|209-1069|TEM-84 [Escherichia coli]; >gb|AJ277414|1-861|TEM-85 [Klebsiella pneumoniae]; >gb|AJ277415|1-861|TEM-86 [Klebsiella pneumoniae]; >gb|AF250872|1-861|TEM-87 [Proteus mirabilis]; >gb|AY027590|113-973|TEM-88 [Klebsiella pneumoniae]; >gb|AY039040|189-1022|TEM-89 [Proteus mirabilis]; >gb|AF351241|90-950|TEM-90 [Escherichia coli]; >gb|AB049569|1-861|TEM-91 [Escherichia coli]; >gb|AF143804|1-861|TEM-92 [Proteus mirabilis]; >gb|AJ318093|1-861|TEM-93 [Escherichia coli]; >gb|AJ318094|1-861|TEM-94 [Escherichia coli]; >gb|AJ308558|182-1042|TEM-95 [Escherichia coli]; >gb|AY092401|1-861|TEM-96 [Escherichia coli]; >gb|AF495873|1-861|TEM-101 [Escherichia coli]; >gb|AY040093|69-929|TEM-102 [Plasmid pWW100]; >gb|AF516719|215-1075|TEM-104 [Klebsiella pneumoniae]; >gb|AF516720|215-1075|TEM-105 [Escherichia coli]; >gb|AY101578|215-1075|TEM-106 [Escherichia coli]; >gb|AY101764|207-1067|TEM-107 [Klebsiella pneumoniae]; >gb|AF506748|39-899|TEM-108 [Salmonella enterica subsp. enterica serovar Typhimurium]; >gb|AY628175|211-1071|TEM-109 [Escherichia coli]; >gb|AY072920|1-861|TEM-110 [Klebsiella pneumoniae]; >gb|AF468003|1-861|TEM-111 [Escherichia coli]; >gb|AY589493|167-1027|TEM-112 [Escherichia coli]; >gb|AY589494|194-1054|TEM-113 [Proteus mirabilis]; >gb|AY589495|182-1042|TEM-114 [Klebsiella aerogenes]; >gb|AF535127|209-1069|TEM-115 [Klebsiella pneumoniae]; >gb|U36911.1|1430-2290|TEM-116 [Staphylococcus aureus]; >gb|AY130282|1-764|TEM-117 [Escherichia coli]; >gb|AY130285|1-785|TEM-118 [Klebsiella oxytoca]; >gb|AY243512|209-1069|TEM-120 [Klebsiella oxytoca]; >gb|AY271267|1-861|TEM-121 [Escherichia coli]; >gb|AY307100|1-861|TEM-122 [Escherichia coli]; >gb|AY327539|1-858|TEM-123 [Proteus mirabilis]; >gb|AY327540|1-858|TEM-124 [Morganella morganii]; >gb|AY628176|76-936|TEM-125 [Escherichia coli]; >gb|AY628199|204-1064|TEM-126 [Escherichia coli]; >gb|AY368236|1-861|TEM-127 [Escherichia coli]; >gb|AY368237|1-861|TEM-128 [Escherichia coli]; >gb|AJ746225|1-861|TEM-129 [Klebsiella oxytoca]; >gb|AJ866988|1-861|TEM-130 [Klebsiella pneumoniae]; >gb|AY436361|132-992|TEM-131 [Salmonella enterica subsp. enterica serovar Typhimurium]; >gb|AY491682|1-861|TEM-132 [Klebsiella pneumoniae]; >gb|AY528425|1-861|TEM-133 [Klebsiella pneumoniae]; >gb|AY574271|1-861|TEM-134 [Citrobacter koseri]; >gb|AJ634602.1|7840-8700|TEM-135 [Salmonella enterica subsp. enterica serovar Typhimurium]; >gb|AY826417|1-861|TEM-136 [Klebsiella pneumoniae]; >gb|AM286274|1-861|TEM-137 [Shigella sonnei]; >gb|AY853593|215-1075|TEM-138 [Salmonella enterica]; >gb|DQ072853|1-861|TEM-139 [Klebsiella pneumoniae]; >gb|AY956335|39-899|TEM-141 [Enterobacter cloacae]; >gb|DQ388882|1-861|TEM-142 [Escherichia coli]; >gb|DQ075245|218-1078|TEM-143 [Escherichia coli]; >gb|AM049399|1-861|TEM-144 [Salmonella enterica subsp. enterica serovar Derby]; >gb|DQ105528|1-861|TEM-145 [Escherichia coli]; >gb|DQ105529|1-861|TEM-146 [Escherichia coli]; >gb|DQ279850|1-861|TEM-147 [Pseudomonas aeruginosa]; >gb|AM087454|209-1069|TEM-148 [Escherichia coli]; >gb|DQ369751|1-861|TEM-149 [Klebsiella aerogenes]; >gb|AM183304|209-1069|TEM-150 [Escherichia coli]; >gb|DQ834729|206-1066|TEM-151 [Escherichia coli]; >gb|DQ834728|206-1066|TEM-152 [Escherichia coli]; >gb|FJ807656|1-861|TEM-154 [Escherichia coli]; >gb|DQ679961|115-975|TEM-155 [Proteus mirabilis]; gb|AM941159|209-1069|TEM-156 [Proteus mirabilis]; >gb|DQ909059|1-861|TEM-157 [Enterobacter cloacae]; >gb|EF534736|214-1074|TEM-158 [Escherichia coli]; >gb|EF136376|1-861|TEM-159 [Proteus mirabilis]; >gb|EF136377|1-861|TEM-160 [Proteus mirabilis]; >gb|EF468463|67-927|TEM-162 [Acinetobacter haemolyticus]; >gb|EU815939|1-861|TEM-163 [Escherichia coli]; >gb|EU274580|215-1075|TEM-164 [Klebsiella pneumoniae]; >gb|FJ197316|1-861|TEM-166 [Escherichia coli]; >gb|FJ360884|214-1074|TEM-167 [Escherichia coli]; >gb|FJ919776|209-1069|TEM-168 [Escherichia coli]; >gb|FJ873740|1-858|TEM-169 [Salmonella enterica subsp. enterica serovar Infantis]; >gb|GQ149347|5270-6130|TEM-171 [Escherichia coli]; >gb|GU550123|145-1005|TEM-176 [Escherichia coli]; >gb|FN652295|1-861|TEM-177 [Proteus mirabilis]; >gb|X97254|154-1011|TEM-178 [Serratia marcescens]; >gb|NG_050218.1|1-1061|TEM-181 [Bacteria]; >gb|HQ529916|111-971|TEM-183 [Klebsiella pneumoniae]; >gb|JN227084|309-1169|TEM-186 [Escherichia coli]; >gb|HM246246|212-1069|TEM-187 [Proteus mirabilis]; >gb|JN211012|214-1074|TEM-188 [Salmonella enterica]; >gb|JN254627|1-861|TEM-189 [Escherichia coli]; >gb|JN416112|1-861|TEM-190 [Escherichia coli]; >gb|KY432484.1|1-861|TEM-191 [Acinetobacter baumannii]; >gb|JF949915|1-754|TEM-192 [Klebsiella pneumoniae]; >gb|JN935135|1-861|TEM-193 [Acinetobacter baumannii]; >gb|JN935136|1-861|TEM-194 [Acinetobacter baumannii]; >gb|JN935137|1-861|TEM-195 [Acinetobacter baumannii]; >gb|HQ877606|1-861|TEM-197 [Klebsiella pneumoniae]; >gb|AB700703|162-1022|TEM-198 [Klebsiella pneumoniae]; >gb|JX050178|1-853|TEM-199 [Proteus mirabilis]; >gb|KC149518|1-861|TEM-153 [Escherichia coli]; >gb|HQ317449|1-861|TEM-182 [Haemophilus parainfluenzae]; >gb|JF795538|1-861|TEM-185 [Escherichia coli]; >gb|FR848831|1-861|TEM-184 [Escherichia coli]; >gb|JQ034306|1-861|TEM-196 [Shigella sonnei]; >gb|JX310327|1-861|TEM-201 [Escherichia coli]; >gb|KC900516|1-858|TEM-205 [Pseudomonas aeruginosa]; >gb|KC783461|1-861|TEM-206 [Escherichia coli]; >gb|KC818234|1-861|TEM-207 [Escherichia coli]; >gb|KC865667|184-1044|TEM-208 [Escherichia coli]; >gb|KF240808|1-861|TEM-209 [Klebsiella pneumoniae]; >gb|KF513179|1-861|TEM-211 [Proteus mirabilis]; >gb|KF663615|1-858|TEM-213 [Pseudomonas aeruginosa]; >gb|KP050491|1-861|TEM-214 [Escherichia coli]; >gb|KP050492|1-861|TEM-215 [Escherichia coli]; >gb|KF944358|1-861|TEM-216 [Escherichia coli]; >gb|HG934763|1-861|TEM-217 [Enterobacter cloacae]; >gb|KM114268|1-861|TEM-219 [Escherichia coli]; >gb|KM998962.1|1-861|TEM-220 [Neisseria gonorrhoeae].

***bla*NDM Gene:**

>gb|FN396876|2407-3219|NDM-1 [Klebsiella pneumoniae]; >gb|JF703135|1-813|NDM-2 [Acinetobacter baumannii]; >gb|JN104597|115-927|NDM-5 [Escherichia coli]; >gb|JQ734687|1-813|NDM-3 [Escherichia coli]; >gb|JQ348841|1-813|NDM-4 [Escherichia coli]; >gb|JN967644|1-813|NDM-6 [Escherichia coli]; >gb|JX262694|1-813|NDM-7 [Escherichia coli]; >gb|AB744718|1-813|NDM-8 [Escherichia coli]; >gb|KC999080|380-1192|NDM-9 [Klebsiella pneumoniae subsp. pneumoniae]; >gb|KF361506|1-813|NDM-10 [Klebsiella pneumoniae subsp. pneumoniae]; >gb|KP265939.1|1-813|NDM-11 [Escherichia coli]; >gb|AB926431|511-1323|NDM-12 [Escherichia coli]; >gb|LC012596|3586-4398|NDM-13 [Escherichia coli]; >gb|KM210086.1|9068-9880|NDM-14 [Acinetobacter lwoffii]; >gb|KP735848.1|1-813|NDM-15 [Escherichia coli]; >gb|KP862821.1|1-813|NDM-16 [Klebsiella pneumoniae]; >gb|KX812714|1-813|NDM-17 [Escherichia coli]; >gb|KY503030.1|1-828|NDM-18 [Escherichia coli]; >gb|MF370080.1|1-813|NDM-19 [Escherichia coli]; >gb|KY654092.1|1-813|NDM-20 [Escherichia coli]; >gb|MG183694.1|1-813|NDM-21 [Escherichia coli]; >gb|MH243357.1|1-813|NDM-22 [Enterobacter cloacae]; >gb|MH450214.1|1-813|NDM-23 [Klebsiella pneumoniae]; >gb|MH450215.1|1-813|NDM-24 [Providencia stuartii]; >gb|MH986670.1|1-813|NDM-25 [Klebsiella pneumoniae]; >gb|MK105832.1|1-813|NDM-27 [Escherichia coli]; >gb|MK425035.1|1-813|NDM-28 [Klebsiella pneumoniae].

***bla*KPC Gene:**

blaKPC-10,"NG_049243.1"; blaKPC-100,"NG_081070.1"; blaKPC-101,"NG_088394.1"; blaKPC-102,"NG_078063.1"; blaKPC-103,"NG_078051.1"; blaKPC-104,"NG_078052.1"; blaKPC-105,"NG_078054.1"; blaKPC-106,"NG_078056.1"; blaKPC-107,"NG_078053.1"; blaKPC-108,"NG_078055.1"; blaKPC-109,"NG_149659.1"; blaKPC-11,"NG_049244.1"; blaKPC-110,"NG_088395.1"; blaKPC-111,"NG_081791.1"; blaKPC-112,"NG_079230.1"; blaKPC-113,"NG_079888.1"; blaKPC-114,"NG_079889.1"; blaKPC-115,"NG_079890.1"; blaKPC-116,"NG_079891.1"; blaKPC-117,"NG_079892.1"; blaKPC-118,"NG_079893.1"; blaKPC-119,"NG_079894.1"; blaKPC-12,"NG_049245.1"; blaKPC-120,"NG_079895.1"; blaKPC-121,"NG_079896.1"; blaKPC-122,"NG_079897.1"; blaKPC-123,"NG_079898.1"; blaKPC-124,"NG_203393.1"; blaKPC-125,"NG_080778.1"; blaKPC-126,"NG_080779.1"; blaKPC-127,"NG_081071.1"; blaKPC-128,"NG_081072.1"; blaKPC-129,"NG_203394.1"; blaKPC-13,"NG_049246.1"; blaKPC-130,"NG_081699.1"; blaKPC-131,"NG_081700.1"; blaKPC-132,"NG_081783.1"; blaKPC-133,"NG_081784.1"; blaKPC-134,"NG_088396.1"; blaKPC-135,"NG_088397.1"; blaKPC-136,"NG_157007.1"; blaKPC-137,"NG_242182.1"; blaKPC-138,"NG_088398.1"; blaKPC-139,"NG_088399.1"; blaKPC-14,"NG_049247.1"; blaKPC-140,"NG_088400.1"; blaKPC-141,"NG_088401.1"; blaKPC-142,"NG_088402.1"; blaKPC-143,"NG_088403.1"; blaKPC-144,"NG_088404.1"; blaKPC-145,"NG_148622.1"; blaKPC-146,"NG_148623.1"; blaKPC-147,"NG_148624.1"; blaKPC-148,"NG_148625.1"; blaKPC-149,"NG_242293.1"; blaKPC-15,"NG_049248.1"; blaKPC-150,"NG_242294.1"; blaKPC-151,"NG_148626.1"; blaKPC-152,"NG_242295.1"; blaKPC-153,"NG_148627.1"; blaKPC-154,"NG_231545.1"; blaKPC-155,"NG_149660.1"; blaKPC-156,"NG_149661.1"; blaKPC-157,"NG_149662.1"; blaKPC-158,"NG_228670.1"; blaKPC-159,"NG_157008.1"; blaKPC-16,"NG_049249.1"; blaKPC-160,"NG_157009.1"; blaKPC-161,"NG_157010.1"; blaKPC-162,"NG_157011.1"; blaKPC-163,"NG_157012.1"; blaKPC-164,"NG_157013.1"; blaKPC-165,"NG_157014.1"; blaKPC-166,"NG_157015.1"; blaKPC-167,"NG_157016.1"; blaKPC-168,"NG_242183.1"; blaKPC-169,"NG_242550.1"; blaKPC-17,"NG_049250.1"; blaKPC-170,"NG_231546.1"; blaKPC-171,"NG_242551.1"; blaKPC-172,"NG_242552.1"; blaKPC-173,"NG_242553.1"; blaKPC-174,"NG_242554.1"; blaKPC-175,"NG_242555.1"; blaKPC-176,"NG_242184.1"; blaKPC-177,"NG_242556.1"; blaKPC-178,"NG_203395.1"; blaKPC-179,"NG_203396.1"; blaKPC-18,"NG_049251.1"; blaKPC-180,"NG_203397.1"; blaKPC-181,"NG_228671.1"; blaKPC-182,"NG_228672.1"; blaKPC-183,"NG_228673.1"; blaKPC-184,"NG_228674.1"; blaKPC-185,"NG_231547.1"; blaKPC-186,"NG_231548.1"; blaKPC-187,"NG_231549.1"; blaKPC-189,"NG_231550.1"; blaKPC-19,"NG_049252.1"; blaKPC-190,"NG_231551.1"; blaKPC-191,"NG_231552.1"; blaKPC-192,"NG_231553.1"; blaKPC-193,"NG_242185.1"; blaKPC-194,"NG_242186.1"; blaKPC-195,"NG_242187.1"; blaKPC-196,"NG_242188.1"; blaKPC-197,"NG_242189.1"; blaKPC-2,"NG_049253.1"; blaKPC-201,"NG_242190.1"; blaKPC-202,"NG_242296.1"; blaKPC-203,"NG_242297.1"; blaKPC-204,"NG_242298.1"; blaKPC-205,"NG_242299.1"; blaKPC-206,"NG_242300.1"; blaKPC-207,"NG_242301.1"; blaKPC-208,"NG_242302.1"; blaKPC-209,"NG_242557.1"; blaKPC-21,"NG_049254.1"; blaKPC-211,"NG_242558.1"; blaKPC-212,"NG_242559.1"; blaKPC-213,"NG_242560.1"; blaKPC-214,"NG_242561.1"; blaKPC-215,"NG_242562.1"; blaKPC-216,"NG_242563.1"; blaKPC-22,"NG_049255.1"; blaKPC-23,"NG_060569.1"; blaKPC-24,"NG_049256.1"; blaKPC-25,"NG_051167.1"; blaKPC-26,"NG_051469.1"; blaKPC-27,"NG_052862.1"; blaKPC-28,"NG_052581.1"; blaKPC-29,"NG_055580.1"; blaKPC-3,"NG_049257.1"; blaKPC-30,"NG_054685.1"; blaKPC-31,"NG_055494.1"; blaKPC-32,"NG_055495.1"; blaKPC-33,"NG_056170.1"; blaKPC-34,"NG_057447.1"; blaKPC-35,"NG_060524.1"; blaKPC-36,"NG_061389.1"; blaKPC-37,"NG_061612.1"; blaKPC-38,"NG_062357.1"; blaKPC-39,"NG_063841.1"; blaKPC-4,"NG_049258.1"; blaKPC-40,"NG_064726.1"; blaKPC-41,"NG_065876.1"; blaKPC-42,"NG_064727.1"; blaKPC-43,"NG_064728.1"; blaKPC-44,"NG_065427.1"; blaKPC-45,"NG_065877.1"; blaKPC-46,"NG_065878.1"; blaKPC-47,"NG_074714.1"; blaKPC-48,"NG_074715.1"; blaKPC-49,"NG_071203.1"; blaKPC-5,"NG_049259.1"; blaKPC-50,"NG_068507.1"; blaKPC-51,"NG_067224.1”; blaKPC-52,"NG_067225.1"; blaKPC-53,"NG_068176.1"; blaKPC-54,"NG_067226.1"; blaKPC-55,"NG_068177.1"; blaKPC-56,"NG_068016.1"; blaKPC-57,"NG_068508.1"; blaKPC-58,"NG_070177.1"; blaKPC-59,"NG_070178.1"; blaKPC-6,"NG_049260.1"; blaKPC-60,"NG_070179.1"; blaKPC-61,"NG_070180.1"; blaKPC-62,"NG_073465.1"; blaKPC-63,"NG_073466.1"; blaKPC-64,"NG_073467.1"; blaKPC-65,"NG_073468.1"; blaKPC-66,"NG_070739.1"; blaKPC-67,"NG_074716.1"; blaKPC-68,"NG_074717.1"; blaKPC-69,"NG_074718.1"; blaKPC-7,"NG_049261.1"; blaKPC-70,"NG_074719.1"; blaKPC-71,"NG_070895.1"; blaKPC-72,"NG_070740.1"; blaKPC-73,"NG_070741.1"; blaKPC-74,"NG_070742.1"; blaKPC-75,"NG_070743.1"; blaKPC-76,"NG_070896.1"; blaKPC-77,"NG_070897.1"; blaKPC-78,"NG_071204.1"; blaKPC-79,"NG_071205.1"; blaKPC-8,"NG_049262.1"; blaKPC-80,"NG_073469.1"; blaKPC-81,"NG_073470.1"; blaKPC-82,"NG_073471.1"; blaKPC-83,"NG_079231.1"; blaKPC-84,"NG_074720.1"; blaKPC-85,"NG_074721.1"; blaKPC-86,"NG_074722.1"; blaKPC-87,"NG_074723.1"; blaKPC-88,"NG_074724.1"; blaKPC-89,"NG_079232.1"; blaKPC-90,"NG_076666.1"; blaKPC-91,"NG_076667.1"; blaKPC-92,"NG_079233.1"; blaKPC-93,"NG_080780.1"; blaKPC-94,"NG_076680.1"; blaKPC-95,"NG_076681.1"; blaKPC-96,"NG_078037.1"; blaKPC-97,"NG_078038.1"; blaKPC-98,"NG_078032.1"; blaKPC-99,"NG_088405.1"

***bla*GES Gene:**

>gb|AF156486|1332-2195|GES-1 [Klebsiella pneumoniae]; >gb|AF326355|1-864|GES-2 [Pseudomonas aeruginosa]; >gb|AB113580|1330-2193|GES-3 [Klebsiella pneumoniae]; >gb|AB116260|1330-2193|GES-4 [Klebsiella pneumoniae]; >gb|AY494717|1-864|GES-5 [Escherichia coli]; >gb|AY494718|1-864|GES-6 [Klebsiella pneumoniae]; >gb|AY260546|4478-5341|GES-7 [Escherichia coli]; >gb|AF329699|373-1236|GES-8 [Pseudomonas aeruginosa]; >gb|AY920928|2690-3553|GES-9 [Pseudomonas aeruginosa]; >gb|FJ820124|1124-1987|GES-10 [uncultured bacterium]; >gb|FJ854362|702-1565|GES-11 [Acinetobacter baumannii]; >gb|FN554543|1-864|GES-12 [Acinetobacter baumannii]; >gb|GU169702|609-1472|GES-13 [Pseudomonas aeruginosa]; >gb|GU207844|1-864|GES-14 [Acinetobacter baumannii]; >gb|GU208678|1-864|GES-15 [Pseudomonas aeruginosa]; >gb|HM173356|512-1375|GES-16 [Serratia marcescens]; >gb|HQ874631|1-864|GES-17 [Escherichia coli]; >gb|JQ028729|1-864|GES-18 [Pseudomonas aeruginosa]; >gb|JN596280|1919-2782|GES-19 [Escherichia coli]; >gb|JN596280|2848-3711|GES-20 [Escherichia coli]; >gb|JQ772478|141-1004|GES-21 [uncultured bacterium]; >gb|JX023441|38-901|GES-22 [Acinetobacter baumannii]; >gb|KF179354|1-861|GES-23 [Pseudomonas aeruginosa]; >gb|AB901141|1-864|GES-24 [Acinetobacter baumannii]; >gb|KP096411|1-864|GES-26 [Pseudomonas aeruginosa].

***bla*CTX-M Gene:**

CTX-M 1.1*like* Subgroup:

>gb|Y10278|1-876|CTX-M-3 [Citrobacter freundii]; >gb|AY044436|1436-2311|CTX-M-15 [Escherichia coli]; >gb|AY080894|4-879|CTX-M-22 [Klebsiella pneumoniae]; >gb|AY238472.1|13-888|CTX-M-33 [Escherichia coli]; >gb|DQ061159|347-1222|CTX-M-42 [Escherichia coli]; >gb|DQ303459|2175-3050|CTX-M-54 [Klebsiella pneumoniae]; >gb|DQ885477|1-876|CTX-M-55 [Escherichia coli]; >gb|EF576988|240-1115|CTX-M-66 [Proteus mirabilis]; >gb|FJ815436|196-1071|CTX-M-71 [Klebsiella pneumoniae]; >gb|AY847148|9-884|CTX-M-72 [Klebsiella pneumoniae]; >gb|DQ256091|99-974|CTX-M-82 [Escherichia coli]; >gb|NZ_SQNR01000043.1|1506-2381|CTX-M-88 [Enterobacteriaceae]; >gb|HQ398214|250-1125|CTX-M-101 [Escherichia coli]; >gb|HG423149|40-915|CTX-M-103 [Escherichia coli]; >gb|GQ351346|1-876|CTX-M-114 [Providencia rettgeri]; >gb|JN227085|352-1227|CTX-M-117 [Escherichia coli]; >gb|KC107824|1-876|CTX-M-139 [Escherichia coli]; >gb|KF240809|1-876|CTX-M-142 [Escherichia coli]; >gb|KJ020573|275-1150|CTX-M-144 [Escherichia coli]; >gb|AJ704396|1-876|CTX-M-96 [Klebsiella pneumoniae]; >gb|EF219134|2126-3001|CTX-M-62 [Klebsiella pneumoniae]; >gb|KM211509|1-876|CTX-M-156 [Klebsiella pneumoniae]; >gb|KM211510|1-876|CTX-M-157 [Klebsiella pneumoniae]; >gb|KP681697.1|1-876|CTX-M-162 [Klebsiella oxytoca]; >gb|KP681698.1|1-876|CTX-M-163 [Escherichia coli]; >gb|KP727571.1|1-876|CTX-M-164 [Proteus mirabilis]; >gb|DQ223685|1-876|CTX-M-52 [Klebsiella pneumoniae]; >gb|EU202673|1-876|CTX-M-80 [Klebsiella pneumoniae]; >gb|KC351754|158-1033|CTX-M-136 [Proteus mirabilis]; >gb|KM211508|1-876|CTX-M-155 [Klebsiella pneumoniae]; >gb|JF966749|158-1033|CTX-M-116 [Proteus mirabilis]; >gb|AY005110|1-846|CTX-M-11 [Klebsiella pneumoniae]; >gb|AJ549244|1-876|CTX-M-28 [Escherichia coli]; >gb|JF274244.1|1-864|CTX-M-107 [Shigella sp. SH219]; >gb|JF274245|1-864|CTX-M-108 [Shigella sp. SH223]; >gb|JF274248.1|1-865|CTX-M-109 [Shigella sp. SH361]; >gb|EU402393|1-876|CTX-M-69 [Escherichia coli]; >gb|EF426798|1-876|CTX-M-79 [Escherichia coli]; >gb|AY267213|1-876|CTX-M-29 [Escherichia coli]; >gb|AY292654|1-876|CTX-M-30 [Citrobacter freundii]; >gb|AF305837|1-876|CTX-M-12 [Klebsiella pneumoniae]; >gb|AM411407|1-876|CTX-M-60 [Klebsiella pneumoniae].

CTX-M 1.2*like* Subgroup:

>gb|X92506|1-876|CTX-M-1 [Escherichia coli]; >gb|AJ557142|1-876|CTX-M-32 [Escherichia coli]; >gb|AB177384|1-876|CTX-M-36 [Escherichia coli]; >gb|EF210159|1-876|CTX-M-58 [Escherichia coli]; >gb|EF219142|11-886|CTX-M-61 [Salmonella enterica subsp. enterica serovar Typhimurium]; >gb|KM211691|1-876|CTX-M-158 [Escherichia coli]; >gb|LN830266.1|1-876|CTX-M-166 [Escherichia coli]; >gb|AF488377|63-938|CTX-M-23 [Escherichia coli]; >gb|AY598759|1-876|CTX-M-10 [Klebsiella pneumoniae]; >gb|AY515297|1-876|CTX-M-34 [Escherichia coli]; >gb|DQ268764|5892-6767|CTX-M-53 [Salmonella enterica subsp. enterica serovar Westhampton]; >gb|AY649755|18-893|CTX-M-37 [Enterobacter cloacae]; >gb|EU177100|5-880|CTX-M-68 [Klebsiella sp. ARS06-441]; >gb|JN790864|239-1114|CTX-M-123 [Escherichia coli]; >gb|JX313020|1-876|CTX-M-132 [Escherichia coli]; >gb|AB284167|226-1101|CTX-M-64 [Shigella sonnei].

CTX-M 2*like* Subgroup:

>gb|DQ125241|1-876|CTX-M-2 [Escherichia coli]; >gb|AJ416344|304-1179|CTX-M-20 [Proteus mirabilis]; >gb|AJ567481|1-876|CTX-M-31 [Providencia sp. 4440]; >gb|AB176534|1-876|CTX-M-35 [Klebsiella oxytoca]; >gb|EF374097|1-876|CTX-M-56 [Escherichia coli]; >gb|DQ408762|1-876|CTX-M-59 [Klebsiella pneumoniae]; >gb|GU127598|1-876|CTX-M-92 [Escherichia coli]; >gb|JN969893.3|2531-3406|CTX-M-131 [Providencia rettgeri]; >gb|KC964871|1-876|CTX-M-141 [Klebsiella pneumoniae]; >gb|DQ102702|1-876|CTX-M-43 [Acinetobacter baumannii]; >gb|D37830|91-966|CTX-M-44 [Escherichia coli]; >gb|KP727572.1|1-876|CTX-M-165 [Klebsiella pneumoniae]; >gb|KJ911020|112-987|CTX-M-115 [Acinetobacter baumannii]; >gb|JQ429324|1-876|CTX-M-124 [Escherichia coli]; >gb|U95364|6-881|CTX-M-5 [Salmonella enterica subsp. enterica serovar Typhimurium]; >gb|AM982520|5549-6424|CTX-M-76 [Kluyvera ascorbata]; >gb|AM982521|1912-2787|CTX-M-77 [Kluyvera ascorbata]; >gb|FN813245|1912-2787|CTX-M-95 [Kluyvera ascorbata]; >gb|Y14156|1-876|CTX-M-4 [Salmonella enterica subsp. enterica serovar Typhimurium]; >gb|AJ005045|1-876|CTX-M-7 [Salmonella enterica subsp. enterica serovar Typhimurium]; >gb|AJ005044|1-876|CTX-M-6 [Salmonella enterica subsp. enterica serovar Typhimurium]; >gb|GQ149243|1-873|CTX-M-74 [Enterobacter cloacae]; >gb|GQ149244|1-873|CTX-M-75 [Providencia stuartii].

CTX-M 8*like* Subgroup :

>gb|AY157676|1-876|CTX-M-26 [Klebsiella pneumoniae]; >gb|AY954516|1-876|CTX-M-39 [Escherichia coli]; >gb|HM167760|1-876|CTX-M-94 [Escherichia coli]; >gb|FR682582|1-876|CTX-M-100 [Escherichia coli]; >gb|DQ023162|1-876|CTX-M-41 [Proteus mirabilis]; >gb|AF518567|2321-3196|CTX-M-25 [Escherichia coli]; >gb|FJ971899|31-906|CTX-M-89 [Proteus mirabilis]; >gb|KP050493.1|1-876|CTX-M-160 [Proteus mirabilis]; >gb|GQ870432|31-906|CTX-M-91 [Proteus mirabilis]; >gb|AM982522|1-876|CTX-M-78 [Kluyvera georgiana]; >gb|KJ461948|4-876|CTX-M-152 [Kluyvera sp. MRB7]; >gb|AY750914.2|207-1079|CTX-M-40 [Escherichia coli]; >gb|AB205197|4-876|CTX-M-63 [Klebsiella pneumoniae]; >gb|AF189721|274-1149|CTX-M-8 [Citrobacter amalonaticus].

CTX-M 9*like* Subgroup:

>gb|AF252622|1741-2616|CTX-M-14 [Escherichia coli]; >gb|AF325134|1-876|CTX-M-19 [Klebsiella pneumoniae]; >gb|AY143430|1-876|CTX-M-24 [Klebsiella pneumoniae]; >gb|AY156923|1-876|CTX-M-27 [Escherichia coli]; >gb|AY822595|23-898|CTX-M-38 [Klebsiella pneumoniae]; >gb|AY847143|83-958|CTX-M-47 [Escherichia coli]; >gb|AY847144|82-957|CTX-M-48 [Klebsiella pneumoniae]; >gb|AY847145|82-957|CTX-M-49 [Klebsiella pneumoniae]; >gb|AY847146|83-958|CTX-M-50 [Klebsiella pneumoniae]; >gb|EF418608|10-885|CTX-M-65 [Escherichia coli]; >gb|EF581888|1-876|CTX-M-67 [Escherichia coli]; >gb|FJ214366|1-876|CTX-M-83 [Salmonella enterica subsp. enterica serovar Derby]; >gb|FJ214367|1-876|CTX-M-84 [Salmonella enterica subsp. enterica serovar Derby]; >gb|FJ214369|1-876|CTX-M-86 [Salmonella enterica subsp. enterica serovar Agona]; >gb|HQ166709|1-876|CTX-M-93 [Escherichia coli]; >gb|HM755448|245-1120|CTX-M-98 [Escherichia coli]; >gb|HM803271|1-876|CTX-M-99 [Klebsiella pneumoniae]; >gb|HQ398215|245-1120|CTX-M-102 [Escherichia coli]; >gb|HQ833652|236-1111|CTX-M-104 [Escherichia coli]; >gb|HQ833651|245-1120|CTX-M-105 [Escherichia coli]; >gb|JF274243|1-876|CTX-M-111 [Shigella sp. SH202]; >gb|JF274246|1-876|CTX-M-112 [Shigella sp. SH257]; >gb|JF274247|1-876|CTX-M-113 [Shigella sp. SH284]; >gb|JN790862|245-1120|CTX-M-121 [Escherichia coli]; >gb|JN790863|233-1108|CTX-M-122 [Escherichia coli]; >gb|JQ724542|175-1050|CTX-M-125 [Enterobacter cloacae]; >gb|JX017364|239-1114|CTX-M-129 [Escherichia coli]; >gb|JX896165|1-876|CTX-M-134 [Escherichia coli]; >gb|KF513180|1-876|CTX-M-147 [Klebsiella pneumoniae]; >gb|KJ020574|245-1120|CTX-M-148 [Escherichia coli]; >gb|FJ214368|1-876|CTX-M-85 [Salmonella enterica subsp. enterica serovar Albany]; >gb|AB976602.1|136-1011|CTX-M-159 [Klebsiella pneumoniae]; >gb|EU545409|81-956|CTX-M-87 [Escherichia coli]; >gb|FJ907381|1-876|CTX-M-90 [Salmonella sp. YLD3]; >gb|AY847147|82-957|CTX-M-46 [Klebsiella pneumoniae]; >gb|AB703103|1-876|CTX-M-126 [Escherichia coli]; >gb|KP128034.1|1-876|CTX-M-161 [Escherichia coli]; >gb|HQ913565|1-870|CTX-M-106 [Escherichia coli]; >gb|AY033516|2836-3711|CTX-M-17 [Klebsiella pneumoniae]; >gb|JX017365|245-1120|CTX-M-130 [Escherichia coli]; >gb|JF274242|1-877|CTX-M-110 [Shigella sp. SH165]; >gb|AF252623|1-876|CTX-M-13 [Klebsiella pneumoniae]; >gb|EU136031|1-876|CTX-M-81 [Klebsiella pneumoniae]; >gb|AF174129|6336-7211|CTX-M-9 [Escherichia coli]; >gb|AY029068|1-876|CTX-M-16 [Escherichia coli]; >gb|DQ211987|1-876|CTX-M-51 [Escherichia coli]; >gb|AJ416346|557-1432|CTX-M-21 [Escherichia coli]; >gb|D89862|112-981|CTX-M-45 [Escherichia coli]; >gb|AB900900|1-876|CTX-M-137 [Escherichia coli].

***bla*OXA Gene:**

OXA 23 *like* Subgroup:

>gb|AY795964|1-822|OXA-23 [Acinetobacter baumannii]; >gb|AF201828.2|116-937|OXA-27 [Acinetobacter baumannii]; >gb|HM488986|1-822|OXA-165 [Acinetobacter baumannii]; >gb|HM488987|1-822|OXA-166 [Acinetobacter baumannii]; >gb|HM488988|1-822|OXA-167 [Acinetobacter baumannii]; >gb|HM488989|1-822|OXA-168 [Acinetobacter baumannii]; >gb|HM488990|1-822|OXA-169 [Acinetobacter baumannii]; >gb|HM488991|1-822|OXA-170 [Acinetobacter baumannii]; >gb|HM488992|1-822|OXA-171 [Acinetobacter baumannii]; >gb|JN638887|1-822|OXA-225 [Acinetobacter baumannii]; >gb|JQ837239|1-822|OXA-239 [Acinetobacter sp. enrichment culture clone 8407]; >gb|KP050485|1-822|OXA-366 [Acinetobacter baumannii]; >gb|KM087842|1-822|OXA-398 [Acinetobacter baumannii]; >gb|AY288523|1-825|OXA-49 [Acinetobacter baumannii]; >gb|EU571228|823-1644|OXA-133 [Acinetobacter radioresistens]; >gb|APQF01000011.1|291558-292379|OXA-103 [Acinetobacter radioresistens DSM 6976 = NBRC 102413 = CIP 103788]; >gb|FJ194494|172-996|OXA-146 [Acinetobacter baumannii]; >gb|AY762325|116-937|OXA-73 [Klebsiella pneumoniae]; >gb|KM433671|1-822|OXA-422 [Acinetobacter baumannii]; >gb|KM433672|1-822|OXA-423 [Acinetobacter baumannii]; >gb|KP144324|1-822|OXA-435 [Acinetobacter baumannii]; >gb|KP727574.1|1-822|OXA-440 [Acinetobacter baumannii]; >gb|KP264124.1|77-898|OXA-482 [Acinetobacter baumannii]; >gb|KP264125.1|84-905|OXA-483 [Acinetobacter baumannii].

OXA 24/40 *like* Subgroup:

>gb|AF509241|1-828|OXA-24 [Acinetobacter baumannii]; >gb|AF201826|22-849|OXA-25 [Acinetobacter baumannii]; >gb|AF201827|22-849|OXA-26 [Acinetobacter baumannii]; >gb|AM991978|1-828|OXA-139 [Acinetobacter baumannii]; >gb|JQ838185|1-828|OXA-207 [Acinetobacter pittii]; >gb|GU199038|1196-2023|OXA-160 [Acinetobacter baumannii]; >gb|GU199039.2|1206-2033|OXA-72 [Acinetobacter baumannii]; >gb|KP410856.1|1-828|OXA-437 [Acinetobacter baumannii].

OXA 48 *like* Subgroup:

>gb|KR401105.1|1-798|OXA-484 [Klebsiella pneumoniae]; >gb|JX423831|2677-3474|OXA-232 [Escherichia coli]; >gb|JN205800|4141-4938|OXA-181 [Klebsiella pneumoniae]; >gb|KP264119.1|1-798|OXA-416 [Shewanella xiamenensis]; >gb|KP410734.1|1-792|OXA-438 [Escherichia coli]; >gb|KP727573.1|1-786|OXA-439 [Escherichia coli]; >gb|HQ700343|1-786|OXA-163 [Enterobacter cloacae]; >gb|JX893517|1-786|OXA-247 [Klebsiella pneumoniae]; >gb|JQ809466|5375-6172|OXA-204 [Klebsiella pneumoniae]; >gb|CP022089.2|2724286-2725083|OXA-252 [Shewanella sp. FDAARGOS_354]; >gb|JN704570|4039-4836|OXA-199 [Shewanella xiamenensis]; >gb|JX438001|1-798|OXA-245 [Klebsiella pneumoniae]; >gb|AY236073|2188-2985|OXA-48 [Klebsiella pneumoniae]; >gb|HM015773|2127-2924|OXA-162 [Klebsiella pneumoniae]; >gb|JX438000|1-798|OXA-244 [Klebsiella pneumoniae]; >gb|KF900153|1-798|OXA-370 [Enterobacter sp. 87F-2]; >NG_055490.1 Klebsiella pneumoniae 1210 pKp1210 blaOXA gene for OXA-48 family class D beta-lactamase OXA-519, complete CDS.

OXA 51 *like* Subgroup:

>gb|KF048915.1|1-825|OXA-342 [Acinetobacter baumannii]; >gb|HQ425493|1-822|OXA-195 [Acinetobacter nosocomialis]; >gb|EU670845|1595-2419|OXA-138 [Acinetobacter nosocomialis]; >gb|HQ425492|1-822|OXA-194 [Acinetobacter nosocomialis]; >gb|HQ425494|1-822|OXA-196 [Acinetobacter nosocomialis]; >gb|HQ425495|1-822|OXA-197 [Acinetobacter nosocomialis]; >gb|HM113561|1-825|OXA-175 [Acinetobacter baumannii]; >gb|HM113562|1-825|OXA-176 [Acinetobacter baumannii]; >gb|HM113563|1-825|OXA-177 [Acinetobacter baumannii]; >gb|EU019536|1-825|OXA-82 [Acinetobacter baumannii]; >gb|HQ734812|1-825|OXA-201 [Acinetobacter baumannii]; >gb|HM113564|1-825|OXA-178 [Acinetobacter baumannii]; >gb|EU029998|576-1400|OXA-115 [Acinetobacter baumannii]; >gb|EU019535|1-825|OXA-80 [Acinetobacter baumannii]; >gb|KF986256.1|41-865|OXA-375 [Acinetobacter baumannii]; >gb|DQ519089|9-833|OXA-95 [Acinetobacter baumannii]; >gb|NWUK01000007.1|12140-12964|OXA-343 [Acinetobacter baumannii]; >gb|HE963770|1-825|OXA-249 [Acinetobacter baumannii]; >gb|EF650034|1-825|OXA-108 [Acinetobacter baumannii]; >gb|EU255295.1|1-825|OXA-126 [Acinetobacter baumannii]; >gb|EU547445|1-825|OXA-130 [Acinetobacter baumannii]; >gb|AM231719|1-825|OXA-90 [Acinetobacter baumannii]; >gb|HQ734811|1-825|OXA-200 [Acinetobacter baumannii]; >gb|KF057032|1-825|OXA-315 [Acinetobacter baumannii]; >gb|KF057033|1-825|OXA-316 [Acinetobacter baumannii]; >gb|DQ519088|9-833|OXA-94 [Acinetobacter baumannii]; >gb|KF048918.1|1-825|OXA-345 [Acinetobacter baumannii]; >gb|DQ392963|9-833|OXA-88 [Acinetobacter baumannii]; >gb|AY750908|1-825|OXA-65 [Acinetobacter baumannii]; >gb|EU547446|1-825|OXA-131 [Acinetobacter baumannii]; >gb|KR872296.1|1-825|OXA-480 [Acinetobacter baumannii]; >gb|KM588353|1-825|OXA-425 [Acinetobacter baumannii]; >gb|APOR01000009.1|341737-342561|OXA-260 [Acinetobacter baumannii NIPH 1362]; >gb|AB781687|1-825|OXA-254 [Acinetobacter baumannii]; >gb|HQ734813|1-825|OXA-202 [Acinetobacter baumannii]; >gb|HM113558|1-825|OXA-172 [Acinetobacter baumannii]; >gb|HM113559|1-825|OXA-173 [Acinetobacter baumannii]; >gb|HM113560|1-825|OXA-174 [Acinetobacter baumannii]; >gb|EF650035|1-825|OXA-109 [Acinetobacter baumannii]; >gb|EU019534|1-825|OXA-79 [Acinetobacter baumannii]; >gb|DQ309277|1-825|OXA-83 [Acinetobacter baumannii]; >gb|DQ309276|1-825|OXA-84 [Acinetobacter baumannii]; >gb|AY949203|1-825|OXA-76 [Acinetobacter baumannii]; >gb|KF048907.1|1-825|OXA-336 [Acinetobacter baumannii]; >gb|NG_050607.1|101-925|OXA-234 [Acinetobacter baumannii]; >gb|AB634250|1-825|OXA-206 [Acinetobacter baumannii]; >gb|EU255296.1|1-825|OXA-127 [Acinetobacter baumannii]; >gb|EF016356.1|1-825|OXA-66 [Acinetobacter baumannii]; >gb|APQY01000006.1|416599-417423|OXA-263 [Acinetobacter baumannii NIPH 329]; >gb|KJ584914.1|1-825|OXA-414 [Acinetobacter baumannii]; >gb|KF057031|1-825|OXA-314 [Acinetobacter baumannii]; >gb|JX025021|1-825|OXA-241 [Acinetobacter baumannii]; >gb|KF057027|1-825|OXA-121 [Acinetobacter baumannii]; >gb|KF057029|1-825|OXA-312 [Acinetobacter baumannii]; >gb|KF057030|1-825|OXA-313 [Acinetobacter baumannii]; >gb|KF986255|17-841|OXA-374 [Acinetobacter baumannii]; >gb|AY750913|1-825|OXA-71 [Acinetobacter baumannii]; >gb|AM231720|1-825|OXA-100 [Acinetobacter baumannii]; >gb|PYSX01000023.1|160179-161003|OXA-337 [Acinetobacter baumannii]; >gb|KJ135342.1|49-873|OXA-390 [Acinetobacter baumannii]; >gb|KM979379.1|15-839|OXA-432 [Acinetobacter baumannii]; >gb|KJ135344|15-839|OXA-381 [Acinetobacter baumannii]; >gb|KJ135345|15-839|OXA-382 [Acinetobacter baumannii]; >gb|KF048911.1|1-825|OXA-339 [Acinetobacter baumannii]; >gb|NGCN01000023.1|65771-66595|OXA-340 [Acinetobacter baumannii]; >gb|EF650032|1-825|OXA-106 [Acinetobacter baumannii]; >gb|NG_049717.1|14-838|OXA-430 [Acinetobacter baumannii]; >gb|CU468230.2|1959578-1960402|OXA-75 [Acinetobacter baumannii SDF]; >gb|KF986263.1|91-915|OXA-384 [Acinetobacter baumannii]; >gb|KJ584917.1|1-825|OXA-408 [Acinetobacter baumannii]; >gb|JX025022|1-825|OXA-242 [Acinetobacter baumannii]; >gb|DQ445683|1-825|OXA-89 [Acinetobacter baumannii]; >gb|EU220744|1-786|OXA-116 [Acinetobacter baumannii]; >gb|GQ853680|1-825|OXA-149 [Acinetobacter baumannii]; >gb|KF057034|1-825|OXA-317 [Acinetobacter baumannii]; >gb|GQ853681|1-825|OXA-150 [Acinetobacter baumannii]; >gb|KP844571.1|1-825|OXA-442 [Acinetobacter baumannii]; >gb|KM588354|1-825|OXA-426 [Acinetobacter baumannii]; >gb|FJ872530|1-825|OXA-144 [Acinetobacter baumannii]; >gb|AY750910|1-825|OXA-68 [Acinetobacter baumannii]; >gb|EU375515|1-825|OXA-128 [Acinetobacter baumannii]; >gb|KF986254|35-859|OXA-386 [Acinetobacter baumannii]; >gb|GQ423625.1|1-825|OXA-117 [Acinetobacter baumannii]; >gb|JX865394.1|1-825|OXA-441 [Acinetobacter baumannii]; >gb|JN603240|1-825|OXA-217 [Acinetobacter baumannii]; >gb|AY862132|1-825|OXA-78 [Acinetobacter baumannii]; >gb|KM979378.1|15-839|OXA-431 [Acinetobacter baumannii]; >gb|DQ519086|1199-2023|OXA-91 [Acinetobacter baumannii]; >gb|AY949202|1-825|OXA-77 [Acinetobacter baumannii]; >gb|AM279652|1-825|OXA-98 [Acinetobacter baumannii]; >gb|KF986259.1|17-841|OXA-378 [Acinetobacter baumannii]; >gb|KJ135343|15-839|OXA-388 [Acinetobacter baumannii]; >gb|KF048913.1|1-825|OXA-341 [Acinetobacter baumannii]; >gb|KJ780078.1|1-825|OXA-400 [Acinetobacter baumannii]; >gb|KF986262.1|28-852|OXA-383 [Acinetobacter baumannii]; >gb|KJ584922.1|1-825|OXA-413 [Acinetobacter baumannii]; >gb|JN248564|1-825|OXA-223 [Acinetobacter baumannii]; >gb|KM979376.1|21-845|OXA-429 [Acinetobacter baumannii]; >gb|KM588352|1-825|OXA-424 [Acinetobacter baumannii]; >gb|DQ519087|39-863|OXA-93 [Acinetobacter baumannii]; >gb|KF986253|16-840|OXA-385 [Acinetobacter baumannii]; >gb|KJ780076.1|1-825|OXA-401 [Acinetobacter baumannii]; >gb|EF653400|435-1259|OXA-113 [Acinetobacter baumannii]; >gb|KF048909|1-825|OXA-338 [Acinetobacter baumannii]; >gb|HQ998857|1-825|OXA-203 [Acinetobacter baumannii]; >gb|KF048917.1|1-825|OXA-344 [Acinetobacter baumannii]; >gb|EF650037|1-825|OXA-111 [Acinetobacter baumannii]; >gb|KJ584916.1|1-825|OXA-407 [Acinetobacter baumannii]; >gb|HM570035|1-825|OXA-179 [Acinetobacter baumannii]; >gb|KF305666.1|1-825|OXA-259 [Acinetobacter baumannii]; >gb|HM570036|1-825|OXA-180 [Acinetobacter baumannii]; >gb|FR865168|1-825|OXA-216 [Acinetobacter baumannii]; >gb|AY750907|1-825|OXA-64 [Acinetobacter baumannii]; >gb|KF885217|1-825|OXA-365 [Acinetobacter baumannii]; >gb|KJ920338.1|1-825|OXA-404 [Acinetobacter baumannii]; >gb|APRA01000005.1|465154-465978|OXA-262 [Acinetobacter baumannii NIPH 67]; >gb|DQ888718|1-825|OXA-99 [Acinetobacter baumannii]; >gb|KJ584920.1|1-825|OXA-411 [Acinetobacter baumannii]; >gb|KF986257|21-845|OXA-376 [Acinetobacter baumannii]; >gb|KF986258|15-839|OXA-377 [Acinetobacter baumannii]; >gb|EU255291.1|1-825|OXA-122 [Acinetobacter baumannii]; >gb|JN215211|1-825|OXA-219 [Acinetobacter baumannii]; >gb|AJ309734|1-825|OXA-51 [Acinetobacter baumannii]; >gb|EU547447|1-825|OXA-132 [Acinetobacter baumannii]; >gb|KJ584918.1|1-828|OXA-409 [Acinetobacter baumannii]; >gb|KJ584915.1|1-825|OXA-406 [Acinetobacter baumannii]; >gb|KF986260|21-845|OXA-379 [Acinetobacter baumannii]; >gb|EU255292.1|1-825|OXA-123 [Acinetobacter baumannii]; >gb|KJ584921.1|1-825|OXA-412 [Acinetobacter baumannii]; >gb|KJ427797|1-825|OXA-391 [Acinetobacter baumannii]; >gb|GQ853679|1-825|OXA-148 [Acinetobacter baumannii]; >gb|AY750912|1-825|OXA-70 [Acinetobacter baumannii]; >gb|EU255294.1|1-825|OXA-125 [Acinetobacter baumannii]; >gb|KJ780077.1|1-825|OXA-402 [Acinetobacter baumannii]; >gb|KF048919.1|1-825|OXA-346 [Acinetobacter baumannii]; >gb|HE963771|1-825|OXA-250 [Acinetobacter baumannii]; >gb|HE963769|1-825|OXA-248 [Acinetobacter baumannii]; >gb|EF650038|1-825|OXA-112 [Acinetobacter baumannii]; >gb|EF650036|1-825|OXA-110 [Acinetobacter baumannii]; >gb|EF650033|1-825|OXA-107 [Acinetobacter baumannii A424]; >gb|DQ335566|1-825|OXA-92 [Acinetobacter baumannii]; >gb|KJ920337.1|1-825|OXA-403 [Acinetobacter baumannii]; >gb|AB871653|11871-12695|OXA-371 [Acinetobacter baumannii]; >gb|CU459141.1|2175317-2176141|OXA-69 [Acinetobacter baumannii AYE]; >gb|APQV01000009.1|266253-267077|OXA-261 [Acinetobacter baumannii NIPH 201]; >gb|KM979380.1|26-850|OXA-433 [Acinetobacter baumannii]; >gb|EU255293.1|1-825|OXA-124 [Acinetobacter baumannii]; >gb|FR853176|1-825|OXA-208 [Acinetobacter baumannii]; >gb|DQ491200|1-825|OXA-67 [Acinetobacter baumannii]; >gb|DQ149247|1-825|OXA-86 [Acinetobacter baumannii]; >gb|DQ348075|1-825|OXA-87 [Acinetobacter baumannii]; >gb|KF986261|45-869|OXA-380 [Acinetobacter baumannii]; >gb|HE963768|1-825|OXA-120 [Acinetobacter baumannii].

**Text S2:** Consensus sequences obtained for the gene groups analyzed.

***bla*SHV Gene consensus:**

------NNNNNNNNTNTTCGCCNGNNTATTATCTCCCTGTTAGCCACCCTGCCGCTGGCGGTA

CACGCCAGCCCGCAGCCGCTTGAGCAAATTAAACNAAGCGAAAGCCAGC---------------------------------------TGTCGGGCNGCGTAGGCATGATAGAAATGGATCTGGCCAGCGGCCGCACGCT

GACCGCCTGGCGCGCCGATGAACGCTTTCCCATGATGAGCACCTTTAAAGTAGTGCTCTGCGGCGCAGTGCTGGCGCGGGTGGATGCCGGTGACGAA------NNNNNNNNTNTTCGCCNGN

NTATTATCTCCCTGTTAGCCACCCTGCCGCTGGCGGTACACGCCAGCCCGCAGCCGCTTGAGCAAATTAAACNAAGCGAAAGCCAGC---------------------------------------TGTCGGGCNGCGTA

GGCATGATAGAAATGGATCTGGCCAGCGGCCGCACGCTGACCGCCTGGCGCGCCGATGAACGCTTTCCCATGATGAGCACCTTTAAAGTAGTGCTCTGCGGCGCAGTGCTGGCGCGGGTGGATGCCGGTGACGAA------NNNNNNNNTNTTCGCCNGNNTATTATCTCCCTGTTAGCCAC

CCTGCCGCTGGCGGTACACGCCAGCCCGCAGCCGCTTGAGCAAATTAAACNAAGCGAAAGCCAGC---------------------------------------TGTCGGGCNGCGTAGGCATGATAGAAATGGATCTG

GCCAGCGGCCGCACGCTGACCGCCTGGCGCGCCGATGAACGCTTTCCCATGATGAGCACCTTTAAAGTAGTGCTCTGCGGCGCAGTGCTGGCGCGGGTGGATGCCGGTGACGAA------N

NNNNNNNTNTTCGCCNGNNTATTATCTCCCTGTTAGCCACCCTGCCGCTGGCGGTACACGCCAGCCCGCAGCCGCTTGAGCAAATTAAACNAAGCGAAAGCCAGC---------------------------------------TGTCGGGCNGCGTAGGCATGATAGAAATGGATCTGGCCAGCGGCCGCACGCTGACC

GCCTGGCGCGCCGATGAACGCTTTCCCATGATGAGCACCTTTAAAGTAGTGCTCTGCGGCGCAGTGCTGGCGCGGGTGGATGCCGGTGACGAA

***bla*TEM Gene consensus:**

----------------------------------------------------------------------------------------------------ATGAGTATTNAACATTTNCGTGTCGCCCTTATTCCCTTTTTTGCGGCATTTTGCNTTCCTGTTTTTGCTCACCCAGAAACGCTGGTGAAAGTAAAAGATGCTGAAGATNAGTTGGGTGCACGAGTGGGTTACATCGANCTGGATCTCAACAGCGGTAAGATCCTTGAGAGTTTTCGCCCCGAAGAACGTTTTCCAATGNTGAGCACTTTTAAAGTTCTGCTATGTGGNGCGGTATTATCCCGTGTTGACGCCGGGCAAGAGCAACTCGGTCGCCGCATACACTATTCTCAGAATGACTTG

***bla*NDM gene consensus:**

ATGGAATTGCCCAATATTATGCACCCGGTCGCGAAGCTGAGCACCGCATTAGCCGCTGCATTGATGCTGAGCGGGTGCATGCCCGGTGAAATCCGCCCGACGATTGGCCAGCAAATGGAAACTGGCGACCAA---------------CGGTTTGGCGATCTGGTTTTCCGCCAGCTCGCACCGAATGTCTGGCAG

CACACTTCCTATCTCGACATGCCGNGTTTCGGGGCAGTCGCTTCCAACGGTTTGATCGTCAGGGATGGCGGCCGCGTGCTGNTGGTCGATACCGCCTGGACCNAT

***bla*KPC Gene consensus:**

NNNNNNNNNNNNNGCCNTCTAGTTCTGCTGTCTTGTCTCTCATGGCCGCTGGCTGGCTTTTCTGCCACCGCGCTGACCAACCTCGTCGCGGAACCATTCGCTAAACTCGAACAGGACTTTGGCGGCTCCATCGGTGTGTACGCGATNGATACCGGNTCAGGCGCAACTGTAAGTTACCGCGCTGAGGAGCGCTTCCCACTGTGCAGCTCATTCAAGGGCTTTCTTGCTGCCGCTGTGCTGGCTCGCAGCCAGCAGCAGGCCGGCTTGCTGGNCACACCCATCCGTTACGGCAAAAATGCGCTGGTTCNGNGGTCACCCATCTCGGAAAAATATCTGACAACAGGCATGACGGTNNNGGAGCTGTCCGCGGCCGCCGTGCAATACAGTGATAACGCCGCCGCCAATTTGTTGCTGAAGGAGTTGGGCGGCCCGGCCNNNCTGACGGCCTTCATGCGCTCTATCGGCGATACCACGTTCCGTCTGGACCGCTGGGAGCTGGAGNTGAACTCCGCNATCCCAGGCGATGCGCGCGATACCTCATCGCCGCGCGCCGTGACGGAAAGCTTACAAAAACTGACACTGGGCTCTGCACTGGCTGCGNCGCAGCGGCAGCAGNTTGTTGATTGGCTAAAGGGAAACACGACCGGCAACCACCGCATCCGCGCGGCGGTGCCGGCAGACTGGGCAGTCGGAGACAAAACCGGAACCTGCGGAGNGTATGNNNNNNCAAATGACTATGCCGTCGTCTGGCCCACTGGGCGCGCACCTATTGTGTTGGCCGTCTACACCCGGGCGCCTAACAAGGATGACAAGNACAGCGAGGCCGTCATCGCCGCTGCGGCTAGACTCGCGCTCGAGGGATTGGGCNNNNNNNNNNNNNNN

***bla*GES Gene consensus:**

ATGCGCTTCATTCACGCNCTATTACTGGCAGNGATCGCTCACTCTGCATATGCNTCGGAAAAATTAACCTTCAAGACCGATCTTGAGAAGCTAGAGCGCGAAAAAGCAGCTCAGATCGGTGTTGCGATCGTCGATCCCCAAGGAGAGATCGTCGCGGGCCACCGAANGGCGCAGCGNTTTGCAATGTGCTCAACGTTCAAGTTTCCGCTAGCCGCGCTGGTCTTTGAAAGAATTGACTCAGGCACCGAGCGGGGGGATCGAAAACTTTCATATGGGCCGGACATGATCGTCNAATGGTCTCCTGCCACGGAGCGGTTTCTAGCATCGGGACACATGACGGTTCTCGAGGCAGCGCAAGCNGCGGTGCAGCTTAGCGACAATGGGGCTACTAACCTCTTACTGAGAGAAATTGGCGGACCTGCTGCAATGACGCAGTATTTTCGTAAAATTGGCGACTCTGTGAGTCGGCTAGACCGGAAAGAGCCGGAGATGNNCGACAACACACCTGGCGACCTCAGAGATACAACTACGCCTATTGCTATGGCACGTACTGTGGCNAAAGTCCTCTATGGCGGCGCACTGACGTCCACCTCGACCCACACCATTGAGAGGTGGCTGATCGGAAACCAAACGGGAGACGCGACNCTACGAGCGGGTTTTCCTAAAGATTGGGTTGTTGGAGAGAAAACTGGTACCTGCGCCAACGGGGNCCGGAACGACATTGGTTTTTTTAAAGCCCAGGAGAGAGATTACGCTGTAGCGGTGTATACAACGGCCCCGAAACTATCGGCCGTAGAACGTGACGAATTAGTTGCCTCTGTCGGTCAAGTTATTACACAACTCATCCTGAGCACGGACAAATAG

***bla*CTX-M Gene:**

CTX-M 1.*1like Consensus*:

NNGGTTAAAAAATCACTGCGNCAGTTCACGCTGATGGCGACGGCANCCGTCACGCTGTTGTTAGGAAGTGTGCCGCTGTATGCGCAAACGGCGGACGTACAGCAAAAACTTGCCGAATTAGAGCGGCAGTCGGGAGGCAGACTGGGNGTGGCATTGATTAACACAGCNGATAATTCGCAAATACTTTATCGTGCTGATGAGCGCTTNGCGATGTGCAGCACCAGTAAAGTGATGGCCGNGGCCGCGGTGCTGAAGAAAAGTGAAAGCGAACCGANTCTGTTAAATCAGCGAGTTGAGATCAAAAAATCTGACCTNGTTAACTATAATCCGATTGCGGAAAAGCACGTCAATGGGACGATGTCACTGGCTGAGCTTAGCGCGGCCGCGCTACAGTACAGCGATAACGTGGCGATGAATAAGCTGATTGCTCACGTTGGCGGCCCGGCTAGCGTCACCGCGTTCGCCCGACAGCTGGGAGACGAAACGTTCCGTCTCGACCGTACCGAGNCGACGTTAAACACCGCCATTCCGGGCGATCCGCGTGATACCACTTCACCTCGGGCAATGGCGCAAACTCTGCGGAATCTGACGCTGGGTAAAGCATTGGGCGACAGCCAACGGGCGCAGCTGGTGACATGGATGAAAGGCAATACCACCGGTGCAGCGAGCATTCAGGCNGGACTGCCTGCTTCCTGGGTTGTGGGGGATAAAACCGGCAGCGGTGNCTATGGCACCACCAACGATATCGCGGTGATNTGGCCAAAAGATCGTGCGCCGCTGATTCTGGTCACTTACTTCACCCAGCCTCAACCTAAGGCAGAAAGCCGTCGCGATNTATTAGCGTCGGCGGCTAAAATCGTCACCNANNNNNNNNNN

CTX-M 1.*2like Consensus*:

ATGGTTAAAAAATCACTGCGNCAGTTCACGCTGATGGCGACGGCAACCGTCACGCTGTTNTTAGGAAGTGTGCCGCTGNATGCGCAAACGGNGGACGTACAGCAAAAACTTGCCGAATTAGAGCNGCAGTCGGGAGGNNGNCTGGGTGTGGCATTGATTAACACNGCNGATAATTCGCAAATACTTTATCGTGCNGATGAGCGNTTTNCNATGTGCAGNACCAGTAAAGTNATGGCNGNNGCNGCGGTGCTNAAGNANAGTGAAANNNAANNGNANCTGNTNAATCAGCNNGTNGAGATCAANNNNNCNGANNTNGTTAACTANAATCCGATTGCNGAAAANCACGTCNANGGNACNATGNCNCTGGNNGANCTNAGCGNGGCCGCGNTNCAGTACAGCGANAANNNNGCNATGAANAANNTGATTNCNCANNTNGGNGGCCCGGNNNGCGTNACNGCNTTNGCCCGNNNGNTNGGNGANGANACGTTNCGTCTNGANCGNACNGANNCNACGNTNAANACNGCCATTCCNGGCGANCCGNGNGANACCACNNCNCCNCGGGCNATGGCGCANACNNTGCGNNANCTNACGCTGGGTNANGCNNTGGGNGANANCCANCGGGCGCAGNTGGTGACNTGGNTNAAAGGCAATACNACCGGNGCNGCNAGNATTCNGGCNGGNNTNCCNNCNTCNTGGNNTGTGGGNGATAANACCGGCAGCNGTGNCTATGGNACCACCAACGATATCGCGGTGATNTGGCCAAAAGATCGTGCGCCGCTGATTCTGGTCANTTACTTCACCCAGCCNCAACCTAAGGCAGAAAGCCGTCGCGATGTATTAGCGTCGGCGGCTAAAATCGTCACCNACGGTTTGTAN

CTX-M 2*like* Consensus:

ATGATGACNCAGAGCATTCGCCGCNNNATGNTNACNGTGATGGCGACNCTNCCCCTGCTNTTTAGCAGCGCNACNCTGCANGCGCAGNCGAACAGCGTGCANCAGCAGCTGGAAGCNCTGGANAAAAGNNNNGGNGGNCGNCTNGGCGTNGCGCTGATTAACACCGCNGATAANNNNCAGATTCTNTANNNNGCNGATGANCGNTTTGCGATGTGCAGNACCAGNAANGTGATGGCGGCNGCGGCGGTGCTNAAACAGAGCGANAGCGATAANCANCTGCTNAANCAGCGCGTNGAAATNANNNNNAGCGANCTGGTNAACTANAANCCNATTGCNGANAAACANGTNAACGGCACNATGACNCTGGCNGANCTNGGCGCNGNNGCNCTGCAGTATAGCGANAANACNGCNATGAANAANCTGATTGCNCATCTGGGNGGNCCNGATAAAGTGACNGCGTTTGCNCGCNNNNTGGGNGATGANACCTTNCGNCTGGANNGNACCGANNCCACNCTNAANANCGCNATTCCNGGCGANCCGCGNGATACCACCACNCCGCTNGCGATGGCGCAGACCCTGAAAAANCTGACNCTGGGNAAAGCGCTGGCGGAAACNCAGCGNGCNCAGNTGGTGACNTGGCTNAANGGCAANACNACCGGNAGCGCGAGCATTCNNGCGGGNCTGCCGAAANNNTGGGNNGTGGGCGATAAAACCGGCAGCGGNGNTTATGGCACCACCAACGATATNGCGNTNATNTGGCCGGAAAACCANGCNCCGCTGGTNCTGGTGACCTANTTTACCCANCCGGANCAGAANGCGGAAAGNCGNCGNGATNTTCTGGCNGCGGCGGCGAAAATNGTNACCCANGGNTTNNNN

CTX-M *8like Consensus*:

NNNATGAGANANNGCGTNANGCGGNNGATNNTAATGACNACNGCCTGTNTTTCGCTGNTGNTGGNNAGTGNGCCGCTGTNTGCNCANGCGAACGANNTTCANCANAAGCTNGCGGCGCTGGAGAAAAGCAGCGGGGGNCGNNTGGGNGTGGCGNTGATTNACACCGCCGATAACNCNCAGACGCTCTACCGCGCCGANGAGCGNTTTGCNATGTGCAGCACCAGTAANGTGATGGCNGNNGCGGCNGTGCTNAAGCAAAGTGAAACGCAAAANNNNNTNNTGAGTCAGNNGGTTGANATTAANNCNTCNGACNTGNTTAACTACAANCCNATNNCNGAAAANCACGTCAANGGCACGATGACNNTNGNGGANNTGANCGCNGCGGCGNTNCAGTACAGCGANAATACNGCCATGAANAAGCTGATTGCCCATCTNGGGGGGCCGGNTAAAGTGACGGCNTTTGCNCGNNNGATTGGNGATNACACNTTCCGGCTCGATCGTACNGAGCCGACGCTCAACACCGCGATCCCCGGCGACCCGCGCGATACCACCACGCCNTTAGCGATGGCGCAGNCNCTNCGCNATCTNACNTTGGGCANTGCCNTNGGTGANACTCAGCGTGCGCANCTGGTNANGTGGCTGAAAGGCAANACCACCGGNGCTGCCAGCATTCNGGCNGGGCTACCCACATCGTGGGTTGTCGGGGATAAAACCGGCAGCGGNGNTTATGGTACGACGAATGANATCGCNGTNATNTGGCCGGAAGGNCGNGCGCCGCTNNTTCTGGTNACTTACTTCACCCANNCNGAGCNGAAGGCAGANANNCGTCGTGACGTNCTCGCNGCTGCCGCNANAATNGTCACCGACGGTTATTAN

CTX-M *9like Consensus*:

ATGGTGACAAAGAGAGTGCAACGGATGATGTTCGCGGCGGCGGCGTGCATTCCGCTGCTGCTGGGCAGCGCGCCGCTTTATGCGCAGACGAGTGCGGTGCAGCAAAAGCTGGCGGCGCTGGAGAAAAGCAGCGGAGGGCGGCTGGGCGTCNCGCTCATCGATACCGCAGATAATACGCAGGTGCTTTATCGCGGTGATGAACGCTTTCCAATGTGCAGTACCAGTAAAGTTATGGCGGNCGCGGCGGTGCTTAAGCAGAGTGAAACGCAAAAGCAGCTGCTTAATCAGCCTGTCGAGATCAAGCCTGCCGATCTGGTTAACTACAATCCGATTGCCGAAAAACACGTCAACGGCACAATGACGCTGGCAGANCTGAGCGCGGCCGCGTTGCAGTACAGCGACAATACCGCCATGAACAAATTGATTGCCCAGCTCGGTGGCCCGGGAGGCGTGACGGCTTTTGCCCGCGCGATCGGCGATGAGACGTTTCGTCTGGATCGCACTGAANCTACGCTGAATACCGCCATTCCCGGCGACCCGAGAGACACCACCACGCCGCGGGCGATGGCNCAGACGTTGCGTCAGCTTACGCTGGGTCATGCGCTGGGCGAAACCCAGCGGGCGCAGTTGGTGACGTGGCTCAAAGGCAATACGACCGGCGCAGCCAGCATTCGGGCCGGCTTACCGACGTCGTGGACTGNNGGTGATAAGACCGGCAGCGGCGNCTACGGCACCACCAATGATATTGCGGTGATCTGGCCGCAGGGTCGTGCGCCGCTGGTTCTGGTGACCTATTTTACCCAGCCGCAACAGAACGCAGAGNGCCGCCGCGATGTGCTGGCTTCAGCGGCGAGAATCATCGCCGAAGGGNTGTNA

***bla*OXA Gene:**

OXA Consensus Subgroup 23 *like*:

ATGAATAAATATTTTACTTGCTATGTGGTTGCTTCTCTTTTTCTTTCTGGTTGTACGGTTCAGCATAATTTAATAAATGAAACCCNGAGTCAGATTGTTCAAGGACATAATCAGGTGATTCATCAATACTTTGATGAAAAAAACACCTCAGGTGTGCTGGTTATTCAAACAGATAAAAAAATTAATCTATATGGTAATGCTCTAAGCCGCGCAAATACAGAATATGTGCCAGCCTCTACATTTAAAATGTTGAATGCCCTGATCGGATTGGAGAACCAGAAAACGGATATTAATGAAATATTTAAATGGAAGGGCGAGAAAAGGTCATTTACCGCTTGGGAAAAAGACATGACACTAGGAGAAGCCATGAAGCTTTCTGCAGTCCCAGTCTATCAGGAACTTGCNNGACGTATCGGTCTTGATCTCATGCAAAAAGAAGTANAACGTATTGNTTTCGGTAATGCTGAAATTGGACAGCAGGTTGANAATTTCTGGTTGNTAGGNCCATTAAAGGTNACGCCTATTCAAGAGGTAGAGTTTGTTTCNCAATTNGCACATACACAGCTTCCATTTAGTGAAAAAGTGCAGGCTAATGTAAAAAATATGCTNCTTNTAGAAGAGAGTAATGGCTACAANATTTTTGGAAAGACTGGTTGG---GCAATGGATATAAAANCACA

AGTGGGCTGGTTGNCCGGCTGGGTTGAGCAGCCAGATGGAAAAATTGTCGCTTTTGCATTAAANATGGAAATGCGGTCAGAAATGCCNGCATCTATACGTAATGAATTATTGATGAAATCATTAAAACAGCTGAATATTATTTAA

OXA Consensus Subgroup 24/40 *like*:

ATGAAAAAATTTATACTTCCTATATTCAGCATTTCTATTCTAGTTTCTCTCAGTGCATGTTCATCTATTAAAACTAAATCTGAAGATAATTTTCATATTTCTTCTCAGCAACATGAAAAAGCTATTAAAAGCTATTTTGATGAAGCTCAAACACAGGGTGTAATTATTATTAAAGAGGGTAAAAATCTTAGCACCTATGGTAATGCTCTTGCACGAGCAAATAAAGAATATGTCCCTGCATCAACATTTAAGATGCTAANTGCTTTAATCGGGCTAGAAAATCATAAAGCAACAACAAATGAGATTTTCAAATGGGATGGTAAAAAAAGAACTTATCCTATGTGGGAGAAAGATATGACTTTAGGTGAGGCAATGGCATTGTCAGCAGTTCCAGTATATCAAGAGCTTGCAAGACGGACTGGCCTAGAGCTAATGCAGAAAGAAGTAAAGCGGGTTAATTTTGGAAATACAAATATTGGAACACAGGTCGATAATTTTTGGTTAGTTGGCCCCCTTAAAATTACACCAGTACAAGAAGTTAATTTTGCCGATGACCTTGCACATAACCGATTACCTTTTAAATTAGAAACTCAAGAAGAAGTTNAAAAAATGCTTCTAATTAAAGAAGTAAATGGTAGTAAGATTTATGCAAAAAGTGGATGGGNAATGGNTGTTACTNCACAGGTAGGTTGGTTGACTGGTTGGGTGGAGCAAGCTAATGGAAAAAAAATCNCCTTTTCGCTCAACTTAGAAATGAAAGAAGGAATGNCTGGTTCTATTCGTAATGAAATTACTTATAAGTNGCTAGAAAATCTTGGAATCATTTAA

OXA Consensus Subgroup 48 *like*:

ATGCGTGTATTAGCCTTATCGGCTGTGTTTTTGGTGGCATCGATTATCGGAATGCCNGCGGTAGCAAAGGAATGGCAAGAAAACAAAAGTTGGAATGCTCACTTTACTGAANATAAATCACAGGGCGTAGTTGNGCTCTGGAATGAGAATAAGCAGCAAGGATTTACCAATAATCTTAAACGGGCGAACCAAGCATTTTTACCCGCATCTACCTTTAAAATTCCCAATAGCTTGATCGCCCTCGATTTGGGCGTGGTTAAGGATGAACACCAAGTCTTTAAGTGGGATGGACANNNGCGNGATATCGCCNCTTGGAATCGNGANCATNANNTAATNACCGCGATGAANTANTCNGTTNTGCCTGTTNATCAANANTTTGCCCGCCAAATTGGNGAGGCACGTATGAGNAANATGCTNCANGCNTTCGATTATGGNAATGAGGANATNTCGGGCAATNTAGNNANTTTNTGGCTNGANGGTGGNATTCGNATTTCGGCNACNNAGCAAATCNNNTTTTTANGNAAGCTGTATCACAANAAGNTNCACGTNTCNGAGCGNAGNCAGCGNATNGTNAAACAAGCCATGCTNACNGANGCNAATGNNGACTATATTATTCGGGCTAAAACNGGNNNNNNNNNNNNNNNCNANNCTAAGATTGGCTGGTGGGTNGGTTGGGTTGANCTNGATGATAATGTGTGGTTTTTTGCGATGAATATGGATATGCCCACATCGGATGGTTTAGGGCTGCGCCAAGCCATCACAAAAGAAGTGCTCAAACAGGANAAAATTATTCCCTAG

OXA Consensus Subgroup 51 *like*:

ATGAACATTNAANCNCTCTTACTTATAACAAGCGCTATTTTTATTTCAGCC---TGCTCACCTT

ATATAGTGNCTGCTAATCCAAATCACAGNGCTTCAAAATCTGATNNNAAAGCAGAGAAAATTAAAAATTTATTTAACGAAGNACACACTACGGGTGTNTTAGTTATCCANCAAGGNCAAACTCAACAAAGCTATGGTAATGATCTTGCTCGTGCTTCGACCGAGTATGTACCTGCTTCGACCTTCAAAATGCTTAATGCTTTGATCGGCCTTGAGCACCATAAGNCAACCACNACAGAAGTATTTAANTGGNANGGNNAAAAAAGGNTNTTCCCAGAATGGGAAAAGNACATGACCCTAGGCGANGCNATGAAAGCTTCCGCTATTCNNGTTTATCAAGATTTAGCTCGTCGTATTGGACTTGANCTCATGTCTAANGAAGTGAAGCGTGTTGGTTATGGCAATGCAGATATCGGTACCCAAGTCGATAATTTTTGGNTNGTGGGTCCTNTAAAAATTACNCCTCAGCAAGAGGCACANTTTGCTTACAAGCTAGCTAATAAAACGCTTCCATTTAGCCNAAAAGTCCAAGATGAAGTGCAATCCATGNTATTCATAGAAGAAAAGAATGGAAANAAAATATACGCAAAAAGTGGTTGGGGATGGGATGTANACCCACAAGTAGGCTGGTTAACTGGATGGGTTGTTCAGCCTCAAGGNAATATTGTAGCNTTCTCCCTTAACTTAGAAATGAAAAAAGGAATACCTAGCTCTGTTCGAAAAGAGATTACTTATAAAAGNTTAGAACAATTAGGTATTTTATAG

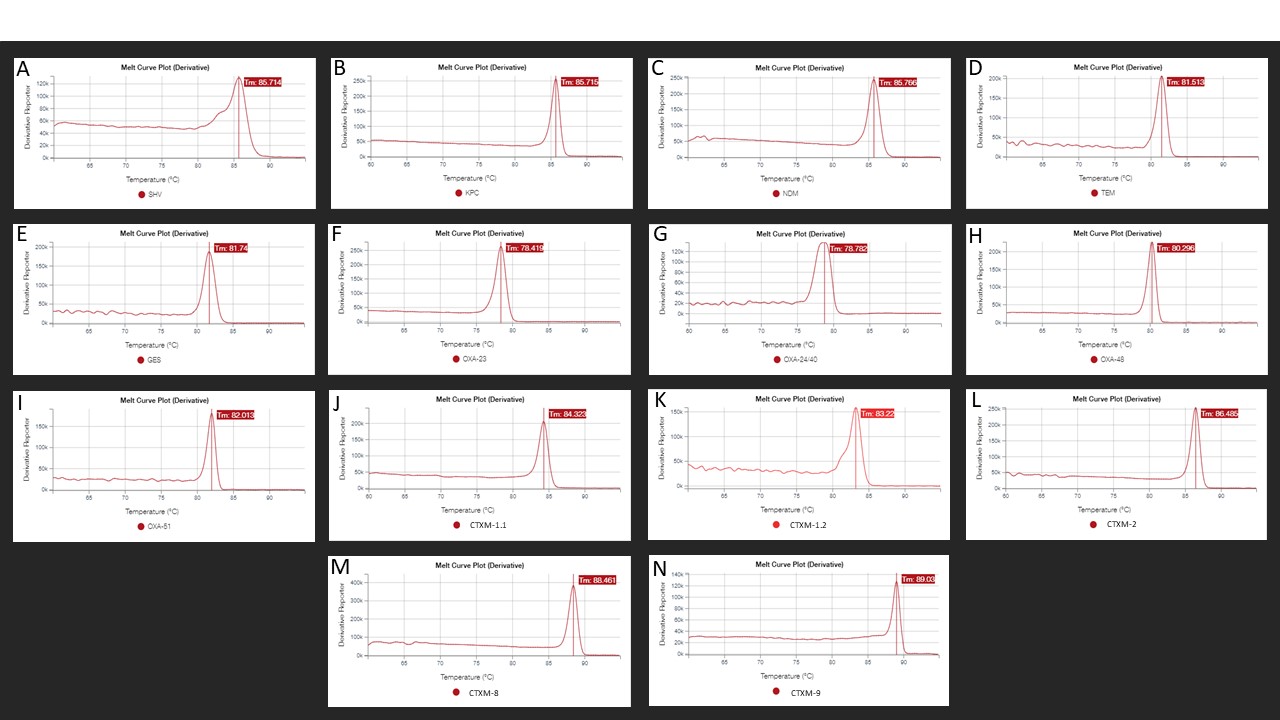

**Figure S1:** Primer’s specificity evaluation through Melting curve: (A) SHV, (B) KPC, (C) NDM, (D) TEM, (E) GES, (F) OXA-23like, (G) OXA-24/40like, (H) OXA-48like, (I) OXA-51like, (J) CTX-M 1.1like, (K) CTX-M 1.2like, (L) CTX-M 2like, (M) CTX-M 8like e (N) CTX-M 9like.

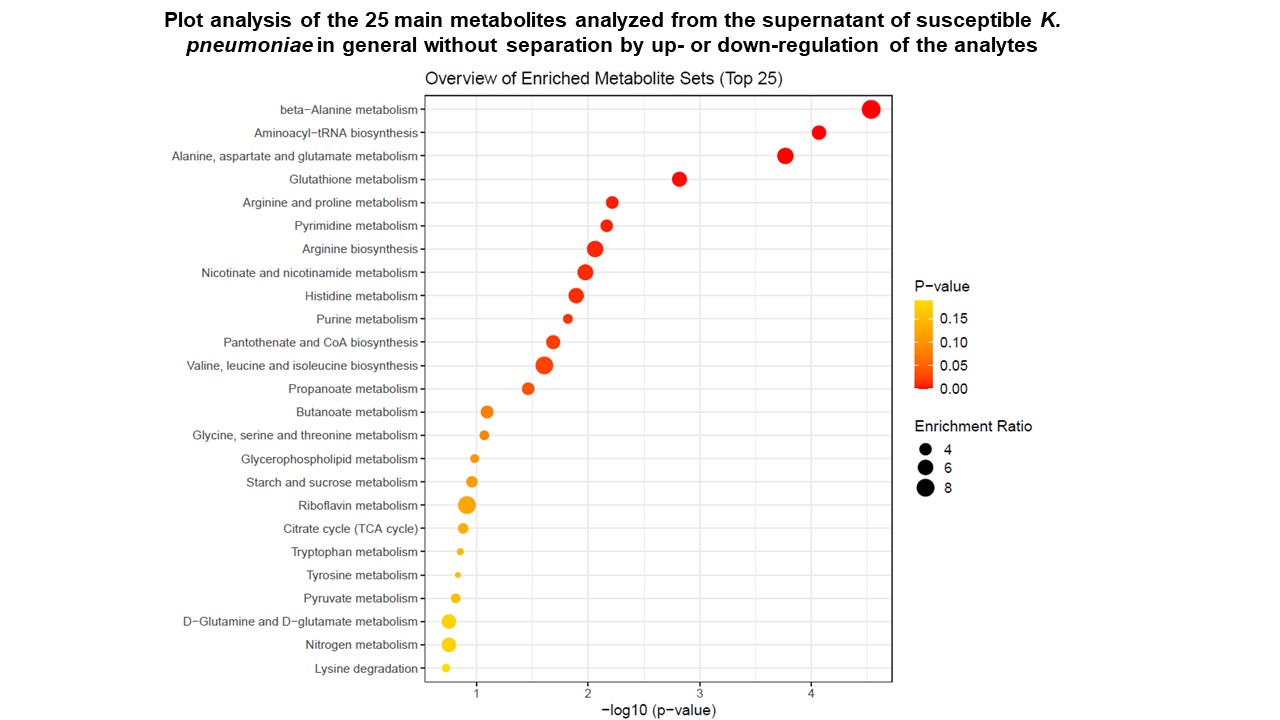

**Figure S2**: Plot analysis of the 25 main metabolites from the supernatant of susceptible *K. pneumoniae* in general without separation by up- or down-regulation of the analytes. Note that in the supernatant of *K. pneumoniae* compared to the culture media, metabolites related to the synthesis of nitrogenous bases, protein synthesis and energy supply to the bacterial cell are significantly increased.

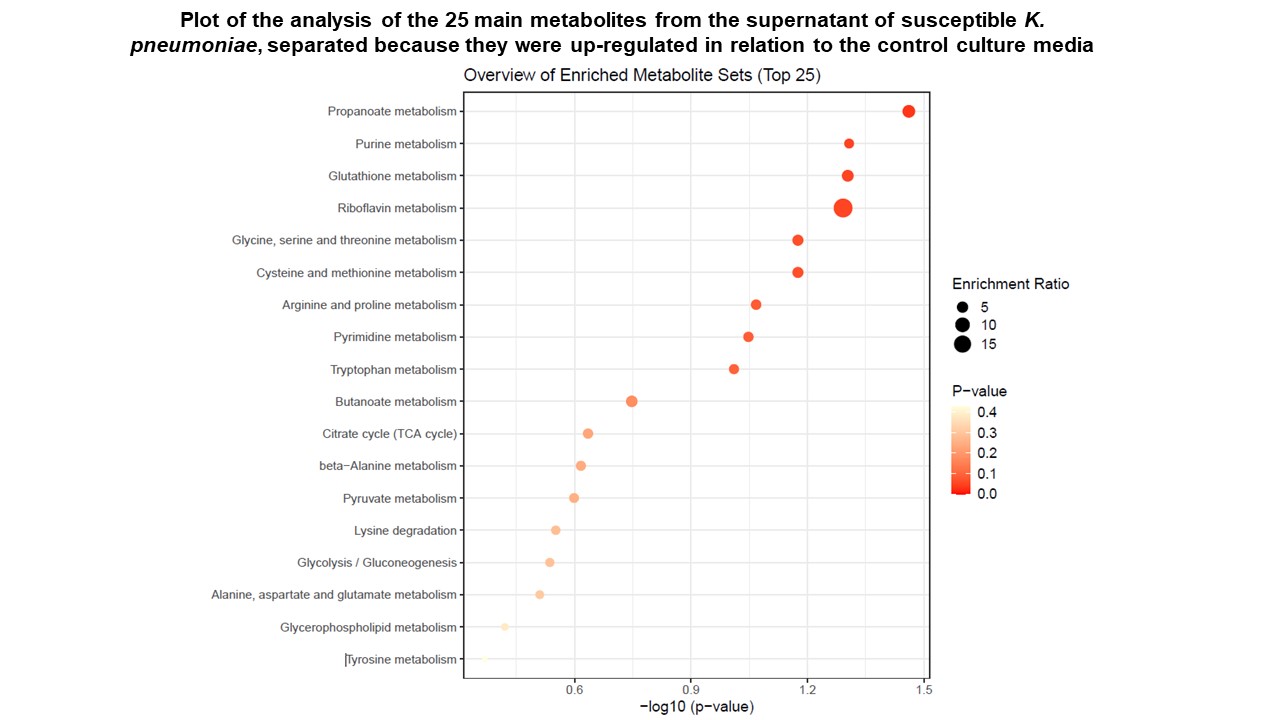

**Figure S3**: Plot of the analysis of the 25 main metabolites from the supernatant of susceptible *K. pneumoniae*, separated because they were up-regulated in relation to the control culture media.

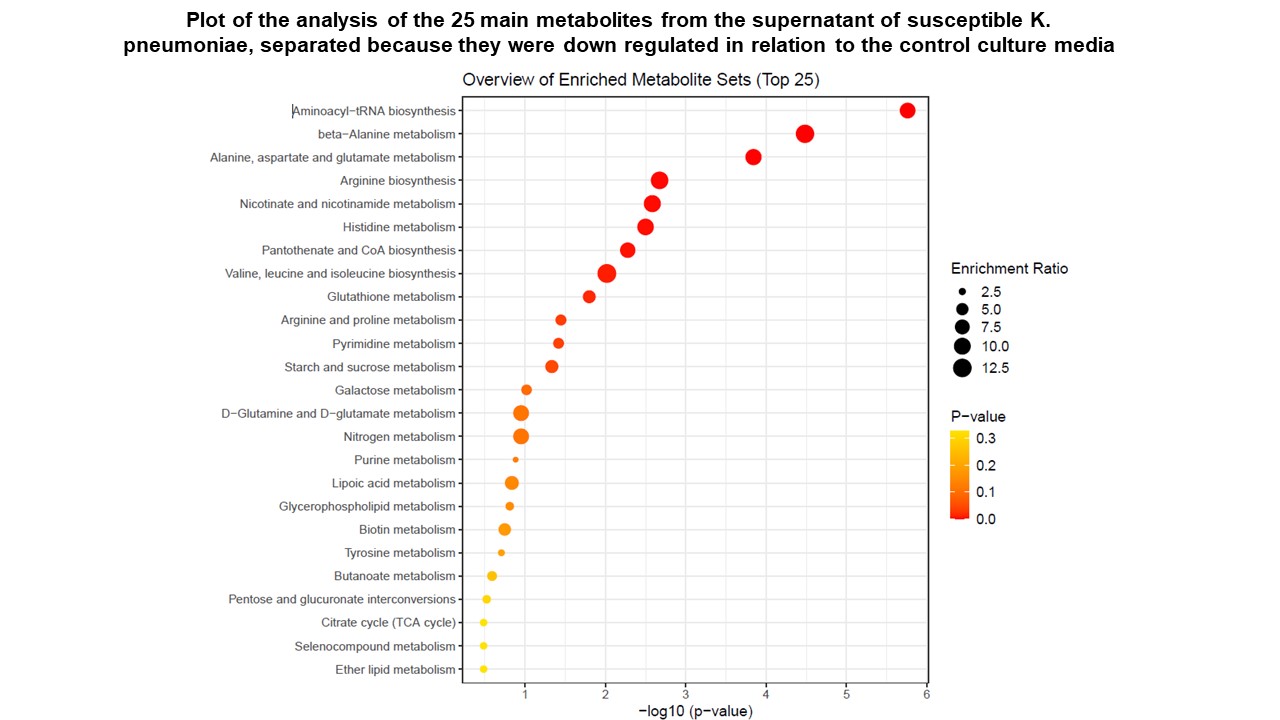

**Figure S4**: Plot of the analysis of the 25 main metabolites from the supernatant of susceptible *K. pneumoniae*, separated because they were down-regulated in relation to the control culture media.

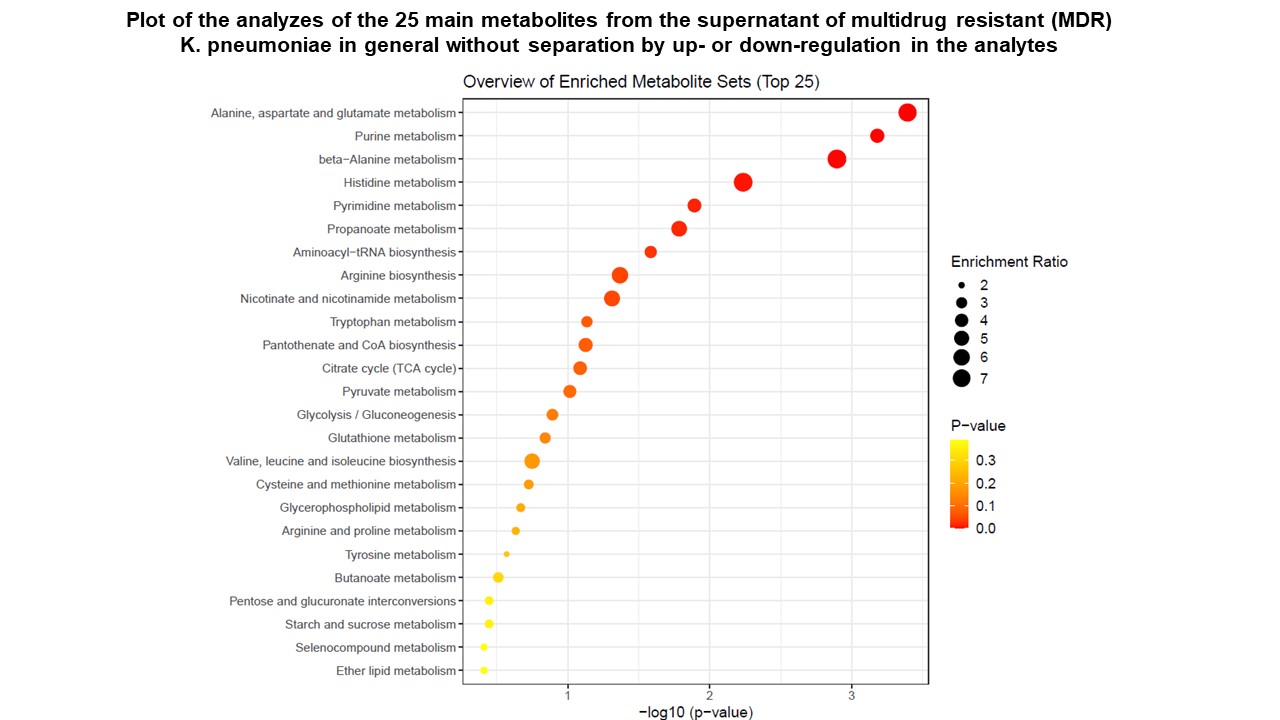

**Figure S5:** Plot of the analyzes of the 25 main metabolites from the supernatant of Multidrug-Resistant (MDR) *K. pneumoniae* in general without separation by up- or down-regulation in the analytes. Note that in the intracellular media of *K. pneumoniae* compared to the culture medium, metabolites related to the synthesis of nitrogenous bases, protein synthesis and energy supply to the bacterial cell are significantly increased.

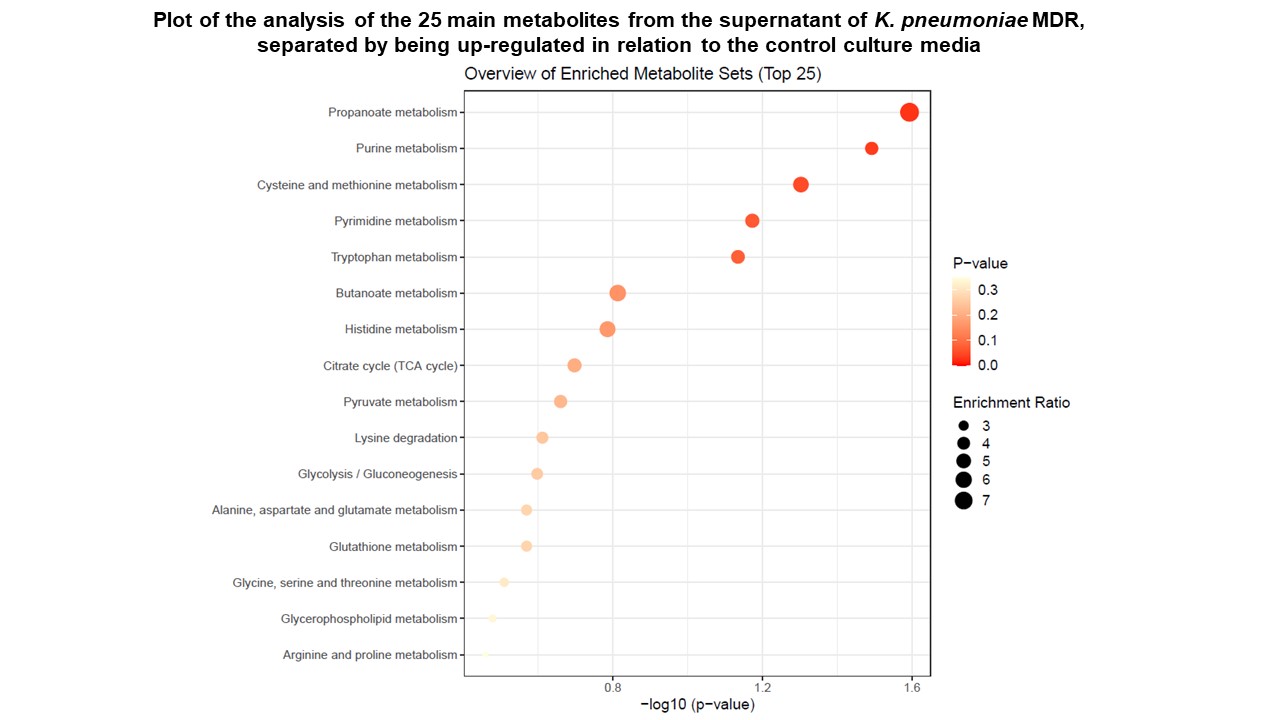

**Figure S6:** Plot of the analysis of the 25 main metabolites from the supernatant of *K. pneumoniae* MDR, separated by being up-regulated in relation to the control culture media.

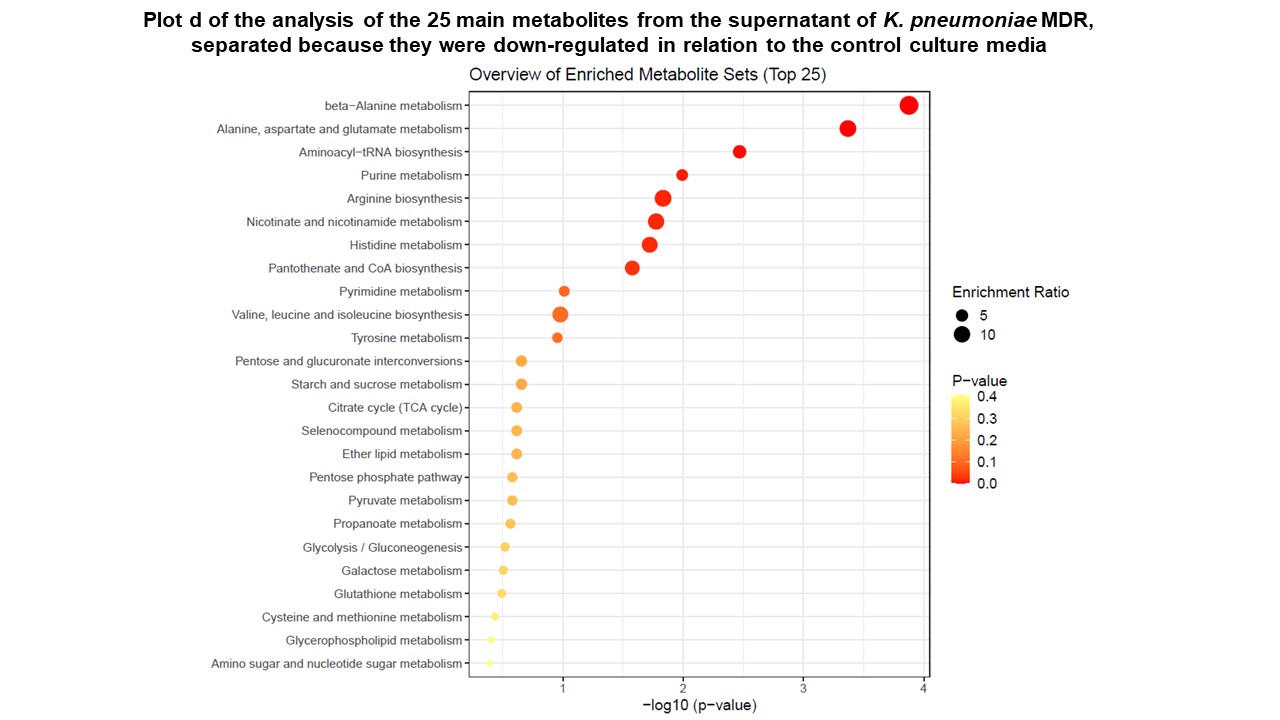

**Figure S7:** Plot d of the analysis of the 25 main metabolites from the supernatant of *K. pneumoniae* MDR, separated because they were down-regulated in relation to the control culture media.
